# Diversity and Scale: Genetic Architecture of 2,068 Traits in the VA Million Veteran Program

**DOI:** 10.1101/2023.06.28.23291975

**Authors:** Anurag Verma, Jennifer E Huffman, Alex Rodriguez, Mitchell Conery, Molei Liu, Yuk-Lam Ho, Youngdae Kim, David A Heise, Lindsay Guare, Vidul Ayakulangara Panickan, Helene Garcon, Franciel Linares, Lauren Costa, Ian Goethert, Ryan Tipton, Jacqueline Honerlaw, Laura Davies, Stacey Whitbourne, Jeremy Cohen, Daniel C Posner, Rahul Sangar, Michael Murray, Xuan Wang, Daniel R Dochtermann, Poornima Devineni, Yunling Shi, Tarak Nath Nandi, Themistocles L Assimes, Charles A Brunette, Robert J Carroll, Royce Clifford, Scott Duvall, Joel Gelernter, Adriana Hung, Sudha K Iyengar, Jacob Joseph, Rachel Kember, Henry Kranzler, Daniel Levey, Shiuh-Wen Luoh, Victoria C Merritt, Cassie Overstreet, Joseph D Deak, Struan F A Grant, Renato Polimanti, Panos Roussos, Yan V Sun, Sanan Venkatesh, Georgios Voloudakis, Amy Justice, Edmon Begoli, Rachel Ramoni, Georgia Tourassi, Saiju Pyarajan, Philip S Tsao, Christopher J O’Donnell, Sumitra Muralidhar, Jennifer Moser, Juan P Casas, Alexander G Bick, Wei Zhou, Tianxi Cai, Benjamin F Voight, Kelly Cho, Michael J Gaziano, Ravi K Madduri, Scott M Damrauer, Katherine P Liao

**Author notes:** **Joint Authorship**. These authors contributed equally to this work (Anurag Verma, Jennifer E Huffman, Alex Rodriguez, Mitchell Conery, Molei Liu). These authors jointly supervised this work (Tianxi Cai, Benjamin F Voight, Kelly Cho, Michael J Gaziano, Ravi K Madduri, Scott M Damrauer, Katherine P Liao). Corresponding authors: Scott M Damrauer.

## Abstract

Genome-wide association studies (GWAS) have underrepresented individuals from non-European populations, impeding progress in characterizing the genetic architecture and consequences of health and disease traits. To address this, we present a population-stratified phenome-wide GWAS followed by a multi-population meta-analysis for 2,068 traits derived from electronic health records of 635,969 participants in the Million Veteran Program (MVP), a longitudinal cohort study of diverse U.S. Veterans genetically similar to the respective African (121,177), Admixed American (59,048), East Asian (6,702), and European (449,042) superpopulations defined by the 1000 Genomes Project. We identified 38,270 independent variants associating with one or more traits at experiment-wide (P < 4.6x10^-11^) significance; fine-mapping 6,318 signals identified from 613 traits to single-variant resolution. Among these, a third (2,069) of the associations were found only among participants genetically similar to non-European reference populations, demonstrating the importance of expanding diversity in genetic studies. Our work provides a comprehensive atlas of phenome-wide genetic associations for future studies dissecting the architecture of complex traits in diverse populations.

**One Sentence Summary:** To address the underrepresentation of non-European individuals in genome-wide association studies (GWAS), we conducted a population-stratified phenome-wide GWAS across 2,068 traits in 635,969 participants from the diverse U.S. Department of Veterans Affairs Million Veteran Program, with results expanding our knowledge of variant-trait associations and highlighting the importance of genetic diversity in understanding the architecture of complex health and disease traits.

## Main text

Among published GWAS, 95% of participants are genetically similar to individuals from European reference populations (*1*). The lack of diversity in genetic research has limited the generalizability of GWAS discoveries (*2*), creating significant concerns about compounding existing disparities in healthcare. This concern can be addressed partly by expanding the representation of diverse populations in large-scale genomic studies. When embraced, such efforts can identify new disease variants (*3*), elucidate disease pathobiology (*4*), and inform treatments that include personalized disease prevention strategies that are broadly applicable to individuals from diverse backgrounds (*5, 6*).

Electronic health record (EHR)-linked biobanks have played a pivotal role in genomic discovery by providing information on the genome and the phenome in large numbers of individuals (*7*–*12*). Historically, genetic association studies have traditionally been performed on cohorts assembled by recruiting participants with specific traits of interest, often from European populations. In contrast, genetic association studies from biobanks with diverse representation enable multi-population analyses that can identify population-specific genetic associations across multiple diseases or traits, assess their clinical significance, reveal pleiotropic effects of genetic variants, and validate previously identified genetic associations across populations (*13, 14*). The growth of increasingly diverse EHR biobanks are poised to begin to address inequities in genetic research (*8, 15–18*).

The Department of Veteran Affairs (VA) Million Veteran Program (MVP) is a longitudinal health, genomic, and precision medicine cohort established in 2011. MVP’s goal is to enroll at least one million Veterans from diverse populations: as of this writing, over 950,000 Veterans have volunteered for the program. The MVP database contains array-based genome-wide genotyping as well as longitudinal EHR data and questionnaire data. Here, we present a multi-population phenome-wide set of GWAS in 635,969 Veterans, of whom 29% are genetically similar to non-European population groups inferred from the 1000 Genomes Project reference panel. We characterize the genetic architecture of 2,068 traits across a comprehensive catalog of phenotypes representing EHR-derived diagnosis codes, clinical laboratory tests, and vital signs, as well as survey responses. We identify independently associated signals using contemporary fine-mapping methods, characterizing pleiotropy across traits and populations, highlighting signals found in non-European population groups (**fig. S1**).

## Results

### Study Design, Population Groups, and Phenotypic Definitions

In this study, we analyzed data from 635,969 participants (MVP Genomics Release 4 (*19*)), aggregated into four population groups based on genetic similarity to the 1000 Genomes Project (*20*) African (AFR, n = 121,177), Admixed Americans (AMR, n = 59,048), East Asian (EAS, n = 6,702), and European (EUR, n = 449,042) superpopulations (**Fig. 1**, **table S1**). After imputation and quality control (QC) filtering, > 44.3M variants (with minor allele count (MAC) > 20) were included for analysis (**Methods**). The frequency and imputation quality scores of single nucleotide polymorphisms (SNPs) among the population groups are provided (**See Data availability**).

**Fig. 1.**
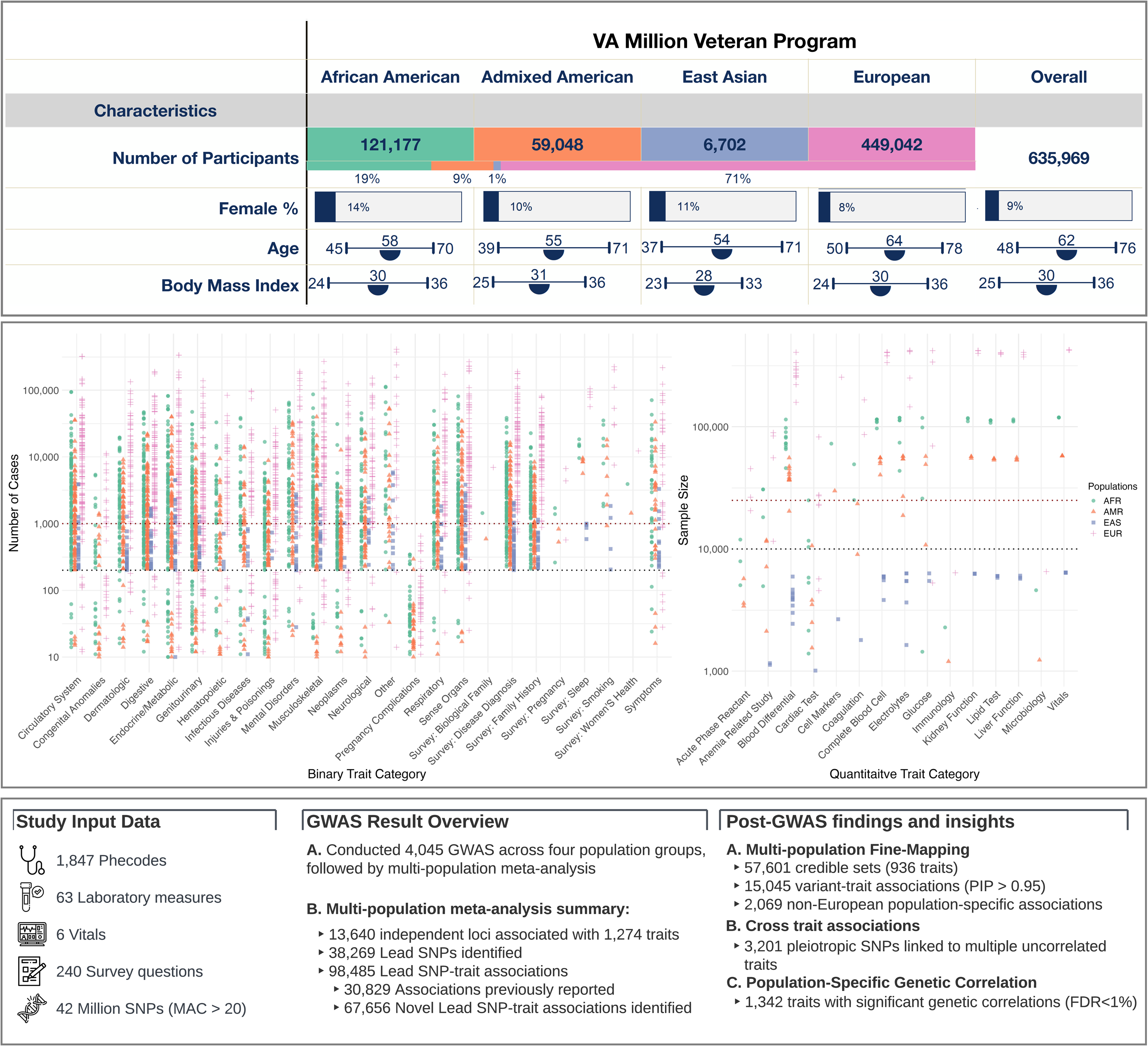
Overview of the study population, genetic association results, and post-GWAS findings. The top panel describes the demographic characteristics of the study population. The middle panel features two scatter plots: the left plot illustrates the number of participants (y-axis) with each binary trait grouped by category(x-axis), and the right plot presents the sample (y-axis) across quantitative traits (x-axis). Distinct colors and shapes represent population groups. The bottom panel is organized into three sections: the left section summarizes the study data, the middle section provides key metrics of GWAS results, such as the count of independent loci and lead SNPs, and the right section briefly outlines the post-GWAS findings.

We extracted phenotypic trait data from the VA EHR, which consisted of diagnosis codes, laboratory measures, and vital signs. Additionally, we included responses to survey questions on health and behavior administered at MVP enrollment. After trait QC, 1,854 binary and 214 quantitative traits were included in the downstream analysis in at least one population group (n=2,068, **Fig. 1, Methods, Supplementary Note**).

### Biobank-scale genomic analysis across populations identifies tens of thousands of trait associations

We next turned to the substantial computational task of generating the >350 billion variant-trait associations across population groups. Unfortunately, the current implementation of the Scalable and Accurate Implementation of GEneralized mixture model (SAIGE) algorithm (*21*) – ideal for our design to address case/control imbalances – was not analytically tractable at this scale of computation and would have required ∼251 compute years to complete. As such, we enhanced the computational efficiency of this algorithm with baseline improvements, implemented a graphics processing unit (GPU) optimization for performing matrix operations, and completed analyses on the US Department of Energy (DOE)’s Oak Ridge Leadership Computing Facility Summit and Andes systems. Using this framework, we conducted a total of 4,045 independent GWAS for traits that met QC criteria in each population group (**table S2**), followed by a multi-population meta-analysis using GWAMA (*22*) for traits present in two or more populations (**Methods**). The actual analysis took 14,286 GPU hours (14 days of wall time), leading to an overall 160-fold reduction in the core hours required.

After multi-population meta-analysis, we identified 98,485 associations (38,269 lead variants at 23,584 loci from 1,274 traits) with a study-wide significance of *P*_<_4.6 × 10^−11^ (**Methods, Fig. 2, and table S3**); 1,096 binary traits (on average, 21 mean associations per trait, **Fig. 2A**) and 178 quantitative traits (on average, 421 mean associations per trait, **Fig. 2B**) exhibited significant associations. The mean genomic inflation factor across all traits was 1.01 (range from 0.85 to 1.19), indicating that the test statistics error rates were relatively controlled (**fig. S2**). Consistent with previous reports (*23*), we noted that the most common variant associations (minor allele frequency (MAF) > 1%) were located in non-coding regions (**fig. S3**); 85.6% (20,158) of variant-binary trait associations (**Fig. 2A, 2B, and tables S4, S5**) and 63.3% (47,496) of variant-quantitative trait associations (**Fig. 2C**, **Fig. 2D, table S4, S5**) have not been previously identified as lead variants or proxies (r^2^ > 0.6 within 500 kb, Methods) in the NHGRI-EBI GWAS (*24*) and Open Target Genetics catalogs (*25*). In fact, 92% (41,979) of the variants associated with quantitative traits and 83% (16,161) with binary traits have not previously been associated with any other trait. Approximately 57% of novel variants had MAF < 1% (**Fig. 2C and 2D**). Further, enrichment analysis to determine whether trait categories were over- or under-represented among novel SNP-trait associations indicated significant over-representation in the 8/22 binary and 6/17 quantitative trait categories (**fig. S4 and table S6**). In particular, this included disease categories such as hematopoietic and infectious disease (OR: 13.08, 95% CI: [7.88-23.49], *P* < 6.58 x 10^-56^; OR: 36.78, 95% CI: [10.12 – 305.07], *P* < 2.03 x 10^-26^), while more well-studied disease categories like neoplasms and circulatory system disorders were less enriched (OR=0.70, 95% CI: [0.62 – 0.79], *P* < 2.87x 10^-09^; OR=0.46, 95% CI: [0.41 – 0.52], *P* < 1.03x 10^-37^) in novel SNP-trait associations.

**Fig. 2.**
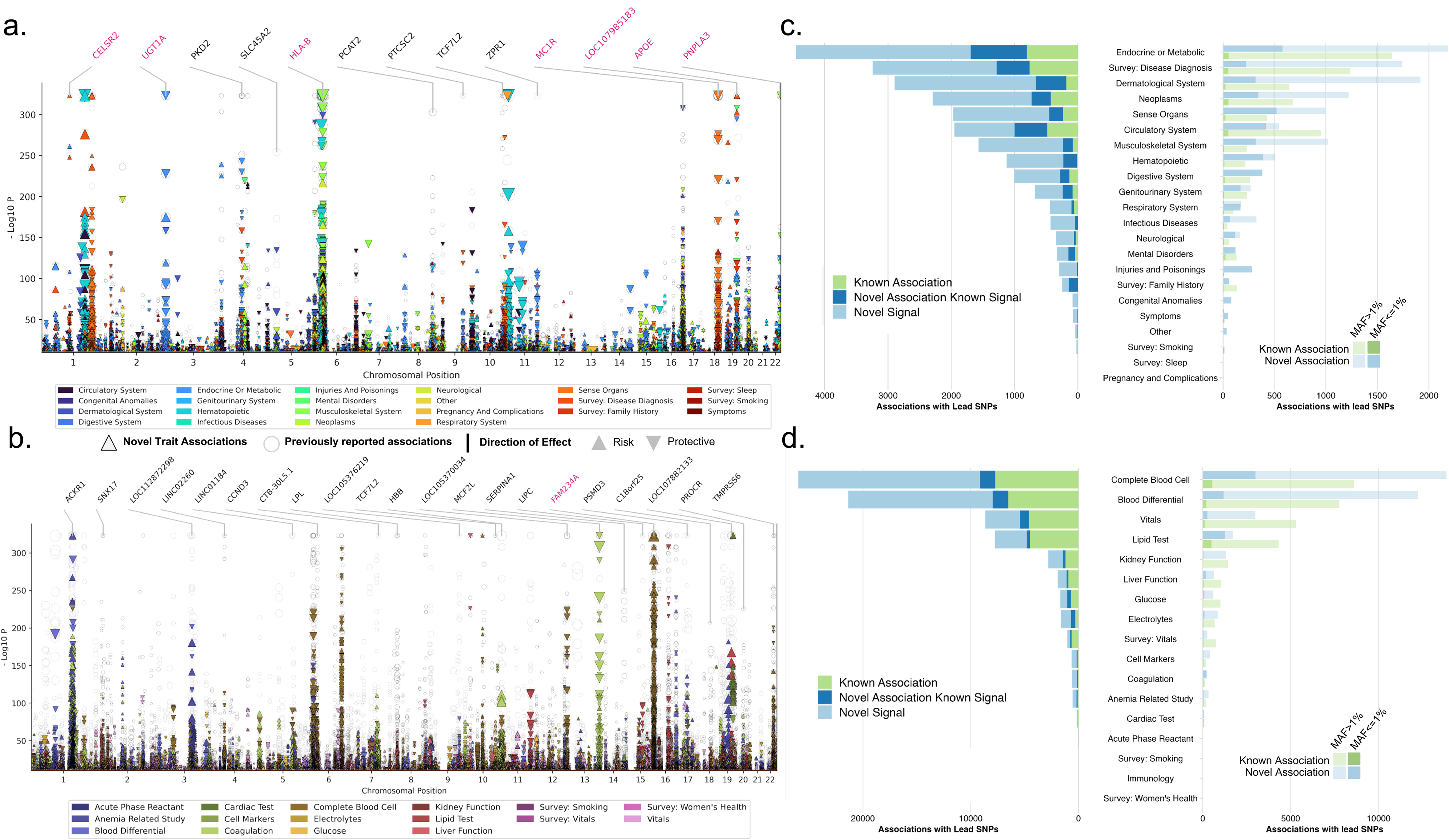
Multi-population genetic associations with traits. Combined multi-trait Manhattan plots and bar plots summarizing 98,485 associations with lead SNPs for binary and quantitative traits (p-value<4.6 x 10^-11^). Manhattan plots for (a) binary traits and (b) quantitative traits display associations across chromosomes (x-axis) and -log10 P values (y-axis). Circles represent previously reported associations, while triangles indicate novel trait associations. The size of the triangles corresponds to the effect size, with upward triangles denoting risk associations and downward triangles signifying protective associations. On the top, gene names indicate previously reported variant-trait associations (in black) and new trait associations (in pink). Stacked bar plot for (c) binary traits and (d) quantitative traits enumerate the number of associations with lead SNPs across different trait categories. The left panel presents the count of known associations (green), novel variants-trait associations (blue), and novel variants (light blue). Trait categories are ordered by the number of lead SNPs in descending order. The right panel is a dodged bar plot highlighting associations with lead SNPs based on their MAF categories: common variants (lower opacity) and low-frequency variants (higher opacity). The distribution of known associations (green) and novel SNPs (light blue) is shown for each trait category.

### Multi-population meta-analysis improves the power to detect significant associations not detected in the EUR population alone

To quantify the suite of discoveries made through expanding representation in genetic analysis, we compared the results of the multi-population meta-analysis to those of the EUR-only GWAS. Over half of the variants analyzed were not included in the EUR population GWAS due to MAF or imputation quality, and a quarter (10M) were only available in AFR (**Supplementary Note**). The inclusion of AFR, AMR, and EAS individuals identified 1,608 additional genomic loci, which were not significant (P > 4.6 x 10^-11^) in the EUR-only analysis (**Table S7**). This led to a total of 3,477 variant-trait associations across 893 traits, 76% of which were with binary traits. The most significant of these results was a rare intronic variant, rs72725854, located near the long non-coding RNA (lncRNA), *PCAT2*, associated with prostate cancer (**table S7**). This single nucleotide polymorphism (SNP) is enriched in African populations (MAF_AFR_ = 0.06, MAF_AMR_ = 0.0068, MAF_EUR_ =0.0006). It has been previously reported to increase the risk of prostate cancer two-fold, aligning with our study findings (*26–28*). We also replicated other previously reported African population-specific associations, such as *ACKR1* for neutropenia and reduced white blood count levels (*29*) and a missense variant, rs73885319 in *APOL1* with kidney-related conditions such as end-stage renal disease (**table S7**) (*30*).

Moreover, we identified 834 variant-trait associations primarily driven by non-EUR populations, as these associations were not even nominally significant in EUR populations (P > 0.05, **table S7**). We discovered an AFR-specific non-coding index variant in *FAM234A* associated with iron deficiency anemias (P_AFR_ = 2.37 x 10^-37^, P_AMR_ = 0.05, P_EUR_=0.42, **table S7**) and hereditary hemolytic anemias (P_AFR_ = 5.32 x 10^-33^, P_AMR_ = 0.28, P_EUR_=0.25, **table S7**). We also identified a novel association of rs3104394 in *MTCO3P1* with alopecia areata reached experiment-wide significance only in AMR (P_AFR_ = 0.01, P_AMR_ = 1.27 x 10^-11^, P_EUR_= 7.66 x 10^-6^). Although there is no information available about the relationship between the *MTCO3P1* gene and alopecia, a cross-sectional analysis of the NIH ’All of Us’ cohort found that alopecia areata is more prevalent in the Hispanic/Latinx community, suggesting potential genetic factors contributing to the development of this condition (*31*).

Lastly, we found 265 variant-trait associations that were significant in AFR or AMR populations and absent in EUR populations. As a positive control, one of the lead variants was rs334 (*HBB*), a well-established variant at increased frequency in African populations due to balancing selection as it is protective against malaria but causes sickle cell anemia risk allele (**table S8).**

### Fine mapping of multi-population associations reveals single-variant credible sets

To create a catalog of putative causal genetic variants at our association signals, we performed signal-based fine-mapping using the Sum of Single Effects model implemented in SuSiE (*32, 33*). For the fine-mapping analysis, we defined 25,953 locus-trait pairs, corresponding to 1,257 traits with one or more study-wide significant variants outside the major histocompatibility complex (MHC) (**fig. S5**). We fine-mapped 99% of these pairs within each population group using exact, in-sample matched linkage disequilibrium (LD) matrices for the trait and identified signals at 22,866 (88%) of the pairs (**fig. S5, Methods**). The 1% of locus-trait pairs that failed to map were primarily due to computational constraints (**table S9**). The fine-mapped signals included 15,045 unique variant-trait pairs (6,318 variants and 613 traits) that mapped with high confidence, i.e. a posterior inclusion probability (PIP) > 0.95 in one or more populations. We merged signals across populations based on their Jaccard similarity indices (*34*) and identified 57,601 multi-population signals across 936 phenotypes (**fig. S5, tables S10-11**); 53,669 (93.1%) of the signals were mapped in a single population including 44,516 (77.3%) that were fine-mapped in only the EUR population (**Fig. 3A**). However, >75% of the signals that were fine mapped in only a single population segregated with one or more unmapped populations (directionally consistent effect estimate and with *P* < 10^-3^) or the underlying GWAS was underpowered to detect a suggestive association in the unmapped population (80% power or less). A larger effective meta-analysis sample size, and thus greater power, was correlated with a larger number of mapped signals (**Fig. 3B**). Among the 15,045 high-confidence pairs, 2,069 variant-trait associations were fine-mapped with high-confidence only in the non-EUR populations (**table S12**). These associations correspond to 974 unique variants and 271 traits (**Fig. 3A**).

**Fig. 3.**
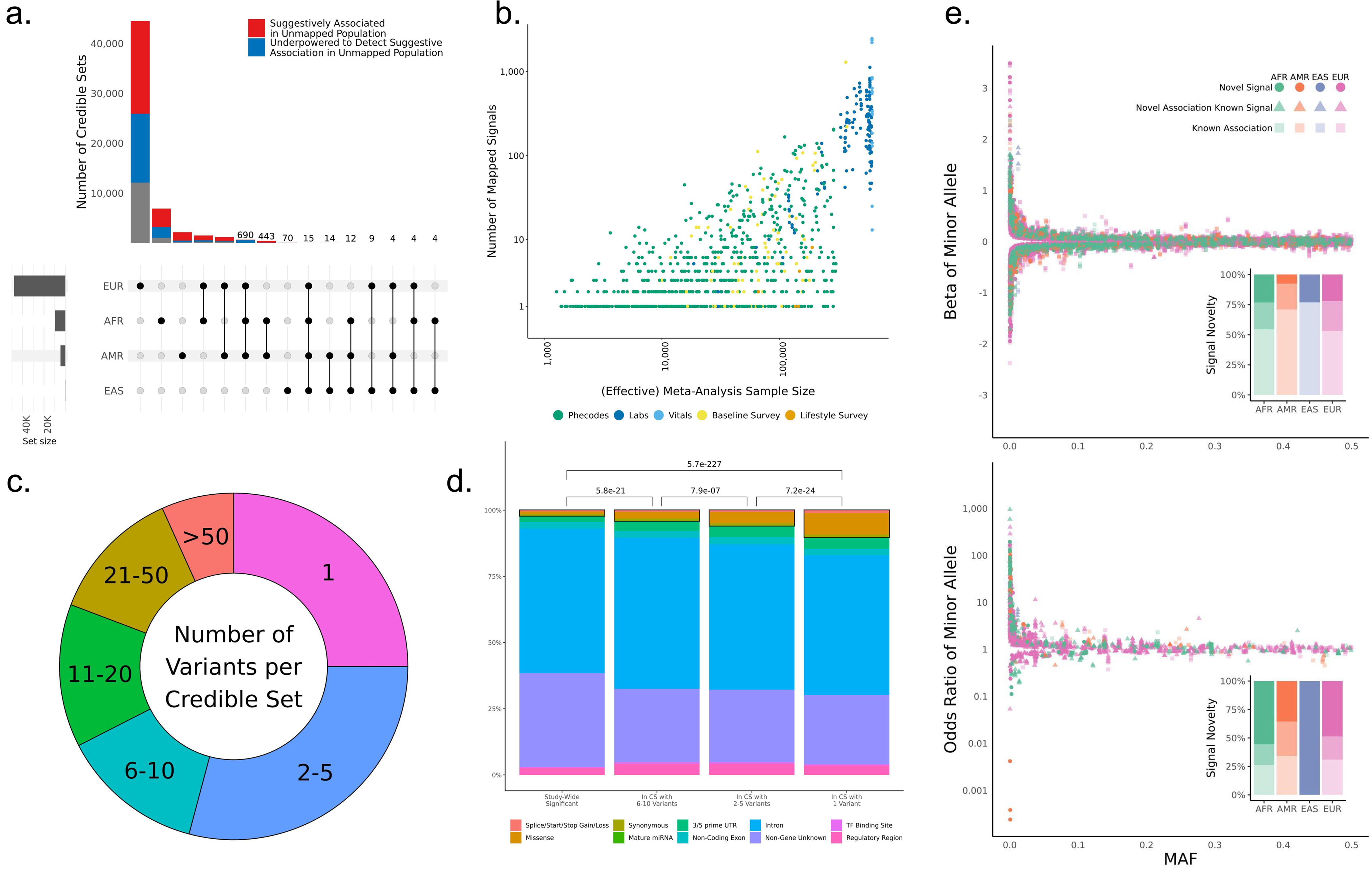
Multi-Population Fine-Mapped Signals. (a) Upset plot of cross-population signal sharing for the 57,601 fine-mapped signals. Red colored-portions of bars represent signals where one or more variants show a suggestive association (*P* < 1x10^-3^) in an unmapped population, and blue colored-portions represent signals where the unmapped populations were underpowered to detect suggestive associations for any of the variants in the merged approximate credible set. Signal counts are displayed above the bars for intersections in which fewer than 1,000 signals were identified. (b) Scatter plot of the number of signals detected per phenotype vs. the meta-analyzed sample size for the phenotype. Effective sample sizes were used for binary phenotypes, and points are colored by the phenotype category. (c) The distribution of merged approximate credible set sizes for the fine-mapped signals. (d) Coding enrichment across increasingly precise fine-mapped signals. Bars are colored by the proportion of each represented by each grouped Variant Effect Predictor (VEP) annotation, and the black boxes illustrate the proportion of each bar attributable to coding variation. P-values reflect the results of pairwise Wilcoxon tests for coding annotation enrichment. (e) Distribution of effect sizes vs. minor allele frequencies for high-confidence (PIP > 0.95) associations fine-mapped in quantitative (top) and binary (bottom) phenotypes. Each point represents a unique high-confidence variant-phenotype-population mapping. Point colors reflect the population in which they are mapped, and their shapes reflect whether they are a phenotype association previously reported in the GWAS catalog (square), a novel phenotype association for a variant previously reported in the catalog (triangle), or a novel SNP and association not previously reported (circle). Inset bar plots reflect the proportions of high-confidence associations in these three categories across the four tested populations.

To quantify the precision of fine mapping for the multi-population results, we conservatively defined an ‘approximate’ credible set for each Jaccard-similarity population-aligned signal as the union of variants in each population-level credible set. Despite this conservative definition, we observed that >54% of the merged signals identified by the fine-mapping pipeline contained ≤5 variants and 14,405 (25%) contained a single variant (**Fig. 3C**). To compare the relative precision of fine mapping between populations, we determined whether there was a difference in the size of our approximate credible sets for signals that mapped in multiple groups. Signals identified in both the AFR and EUR populations generally had slightly but significantly smaller sets in the AFR mapping than in the EUR mapping (Wilcoxon signed-rank *P* = 2.26 × 10^-10^; **fig. S6**). In contrast, our approximate credible sets in AMR mapping were larger than their AFR (*P* = 1.30 × 10^-84^, **fig. S6**) and EUR counterparts (*P* = 7.36 × 10^-162^, **fig. S6**). Overall, we observed an enrichment of annotations in coding regions for variants in signals that fine-mapped to fewer than ten variants, which increased as a function of fine-mapping precision (**Fig. 3D, table S13**). However, even among the single-variant signals, ∼90% of the fine-mapped variants were in non-coding regions. Among the non-coding fine-mapped variants, we observed a slight enrichment of higher functional prediction scores from RegulomeDB (*35*) relative to all significantly-associated variants (**fig. S7**).

We next analyzed the distribution of effect sizes and allele frequencies for lead variants and fine-mapped signals for the 15,822 variant-trait-population combinations with high confidence (PIP > 0.95) fine-mapped signals. Consistent with previous reports (*36, 37*), we observed an inverse relationship across all four population groups between the minor allele frequency of a variant and its effect size for both lead variants (**fig. S8**) and high-confidence signals (**Fig. 3E**). For the high-confidence signals, we additionally examined the relationship between frequencies and effect sizes for alleles derived in the human lineage since the last common ancestor of chimpanzees and bonobos (**Fig. S8**). Because 86.9% of derived alleles were the minor allele, it was not surprising that we saw strong effects for variants with allele frequencies close to 0. Large effect sizes were also observed for several variants with whose derived allele was high-frequency, some of which map to previously reported targets of positive selection in human populations (*38–40*). We observed this relationship between allele frequency and effect size for both newly observed variant-trait associations and those previously reported in the GWAS Catalog (*24*), with similar relative proportions in the three well-powered populations (AFR, AMR, EUR). Interestingly, the distribution of effects in binary and quantitative phenotypes was different. Although it was equally common for minor and derived alleles at high-confidence signals to associate with an increase or decrease in a quantitative trait, e.g., higher white blood cell count (WBC) or lower WBC, (49.6% of minor and derived alleles were associated with a higher value of the quantitative trait), the majority, (71%) of these alleles conveyed increased risk for binary traits. The increased-risk effect among minor alleles was also observed for lead variants; however, only 64% of lead-SNP minor alleles increased the risk (**fig. S8**).

### Cross-trait genetic architecture identifies pleiotropic loci

Next, we investigated the associations of high-confidence fine-mapped variants (PIP > 0.95) with two or more traits. Filtering the 15,045 high-confidence fine-mapped variant-trait pairs, fine-mapped above to include only traits with a phenotypic correlation coefficient > |0.2| resulted in 3,955 variant-trait pairs across 3,201 variants and 377 traits (**table S14**, **fig. S9**). Among these, 87 variants were associated with three or more traits resulting in 360 associations (**Fig. 4**). Although a total of 172 out of these 360 associations reported were previously observed in prior studies (*24, 25*), we detected 188 novel variants associated with multiple traits across 30 trait categories (**Fig. 4**). In particular, the missense variant rs429358 in the *APOE* gene is linked to 24 different traits, including those previously identified conditions such as macular degeneration, abdominal aortic aneurysm, dementia, Alzheimer’s disease, and HDL levels. We observed new associations between this variant and chronic liver disease and cirrhosis.

**Fig. 4.**
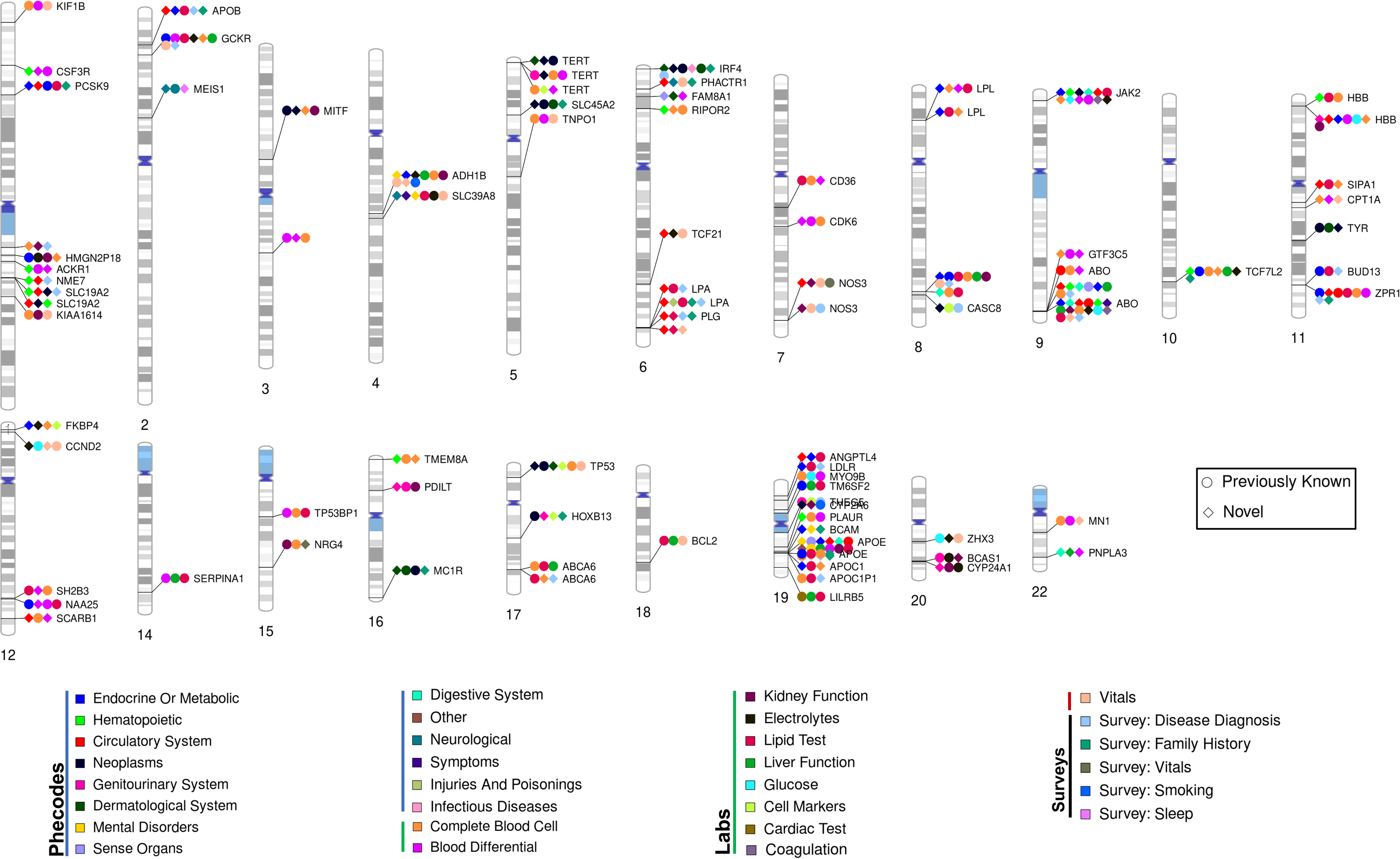
Fine-mapped cross-trait associations. Chromosome ideogram illustrating high-confidence cross-trait associations (PIP > 0.95) between genetic variants and independent traits. Independence was determined using a phenotypic correlation coefficient threshold of 0.2 and a stepwise selection process accounting for the largest PIP value, meta-analysis p-value, population-specific p-values, effect size, and trait specificity (e.g., gender-specific traits). The ideogram highlights genomic regions associated with three or more traits, including both previously reported associations and novel trait associations discovered in the present study. Colors represent distinct trait categories, while shapes distinguish between previously reported (circle) and novel trait associations (triangle). Each data point on the ideogram indicates that at least one trait within the respective category is associated with the genomic region.

### Population-specific heritability and genetic correlation patterns for complex traits

To characterize phenotypic variation attributable to common genetic variants across the four major population groups, SNP heritability was calculated using linkage disequilibrium score regression (LDSC) with population-specific GWAS results and in-sample LD reference panels (**Supplementary Note**). This analysis identified significant (*P* < 9 × 10^-6^) SNP heritability for 233 traits (*n =* 1,525, mean h^2^ = 20.45%) in the AFR, 199 traits (*n =* 1,226, mean h^2^ = 22.12%) in the AMR, three traits (*n =* 353, mean h^2^ = 50.85%) in the EAS, and 816 traits (*n =* 1,898, mean h^2^ = 12.22%) in the EUR groups (**fig. S10, table S15**). Height was the most heritable trait across all four population groups, consistent with a previous report (*41*).

We then examined the genetic correlation between pairs of traits with significant heritability within each population group using LDSC and MVP-derived LD reference panels. We identified significant correlations (false discovery rate < 1%) in all populations: 6,424 pairs (671 traits) in AFR, 3,031 pairs (494 traits) in AMR, and 154,503 pairs (1,342 traits) in EUR populations. Although there was considerable overlap in trait pairs across populations, 1,267 unique trait pairs were identified among non-EUR participants. Classifying variables according to genetic correlations and seeking groups with robust correlations (r_g_ > 0.60) revealed numerous clusters of attributes with shared genetic influences (**table S16)**. Despite significant correlations, no trait exhibited an opposite genetic correlation between the two population groups. However, genetic correlations for many trait pairs varied by population. For instance, mean BMI and type 2 diabetes had a genetic correlation of 0.47 (0.41-0.52) in the EUR group, while only 0.28 (0.18-0.39) in the AFR group.

We finally analyzed the genetic correlation of each trait between the population groups using Popcorn (*42*). With EUR as the reference population, 212 traits exhibited a significant genetic correlation between EUR and AFR (*P*<3.6 x 10^-5^, 0.05 / 1380 traits), 13 between EUR and AMR (*P*<5.61 x 10^-5^, 0.05 / 890 traits), and six between EUR and EAS (*P*<1.4 x 10^-4^, 0.05 / 336 traits, **table S17**). Specifically, we found the strongest genetic correlation for height (p_gi_= 0.66) among quantitative traits and type 2 diabetes (p_gi_= 0.65) among binary traits between EUR and AFR populations. These correlations were not significant when compared with AMR and EAS, likely due to low sample sizes in those populations (**table S17**). We also noted that some traits, including skin cancer (p_gi_ = 0.05), anemia of chronic disease (p_gi_ =0.08), iron levels (p_gi_ = 0.18), and white blood cell counts (p_gi_ = 0.20), had weaker correlations between different populations, in keeping with population-specific variant-trait associations we identified above. Overall, the large number of observed genetic correlations between traits across populations underscores the role of shared genetic factors contributing to trait variance across our population groups.

### Fine-mapped variant-trait associations specific to non-EUR populations

Of the 25,953 high-confidence variant-trait pairs identified by fine-mapping, 2,069 (974 unique variants and 271 phenotypes) were unique in the non-EUR population groups (**table S12**). Although most of the signals were from low frequency or rare variants, 15 novel signals (10 AFR, 3 AMR, 2 EAS) occurred in coding variants and had a MAF>0.05. Among these was a missense variant, rs73382631, associated with lower WBC and neutrophil counts in the AFR population (MAF_AFR_ = 0.10, MAF_AMR_ = 0.01, not present in EAS, EUR). Another example was a missense coding variant in *ABCG2* (rs35965584, MAF_AFR_ = 0.002, not present in AMR, EAS, EUR), for which our analysis identified a novel association with gout. Previous reports have identified an association between *ABCG2* and hyperuricemia (*43*) and susceptibility to gout (*44*), with another known *ABCG2* missense variant of the *ABCG2* gene (rs2231142) (*45*). In MVP, the previously identified missense variant, rs2231142, was within the 95% credible set of a distinct gout signal (n=8 variants) mapped in EAS and EUR but was not in linkage disequilibrium with rs35965584 (r^2^ = 0.0001).

We noted that the majority of population-specific signals were in non-coding regions. To gather insights into these variants, we used functional prediction scores from RegulomeDB (**table S12**), identifying 43 previously known associations and 20 novel associations with SNPs that had strong evidence of being regulatory (RegulomeDB score>0.9). The previously reported loci were associated with factors such as hemoglobin A1c, cholesterol measures, heart rate, red blood cell count, and type 2 diabetes (*46, 47*). All other newly identified associations were related to quantitative traits, such as rs574674363 and lower high-density lipoprotein (HDL) cholesterol levels.

Heterogeneity of effect size was screened across common signals (MAF > 0.05) at 1,888 fine-mapped loci (representing 1,329 separate traits) with overlapping credible sets in multiple populations. This identified 16 heterogeneous variant-trait associations when comparing the AFR to the EUR population and 11 when comparing the AMR to the EUR population (**table S18**). Focusing on coding variants that mapped to the same trait with high confidence (PIP > 0.95) in multiple populations, we observed six associations with significant heterogeneity in effect size between the estimates in the AFR and the EUR populations, and 2 when comparing the AMR to the EUR populations (**table S19**). All variant-trait pairs had the same direction of effect. The majority of differences across signals mapped to rs429358 (MAF_AFR_: AFR= 0.22, MAF_AMR_=0.12, MAF_EUR_=0.14), the coding variant tag for *APOE-e4* associated with a 30% lower risk of dementia in the AFR compared to the EUR population (*48*); there was no heterogeneity in the effect estimate between the AMR and the EUR population. The EAS population was underpowered for this heterogeneity analysis (**fig. S11**).

## Discussion

Here, we present a comprehensive, phenome-wide atlas of GWAS analyses in the largest multi-population biobank to date. Analyzing approximately 44.3 million variants across 2,068 traits in 635,969 US Veterans representing four diverse populations, we identified 98,485 independent variant-trait associations. The summary statistics from this effort comprise a freely accessible resource for the community to dissect the genetic basis of health and disease and foundational data to support discovery, bridging the knowledge gap in the underrepresented populations. Additionally, we highlight the essential role of efficient software operating at exascale supercomputing capabilities to perform analyses at this scale, made possible through a collaboration between the VA and DOE.

Using fine-mapping with SuSiE, we identified 57,601 independent signals across 936 of the traits, including 14,405 signals that were fine-mapped to a single variant, enriched for annotations (e.g., coding variants) that *a priori* are likely causal. Despite this, 90.8% of approximate credible sets that mapped to a single variant carried only “‘non-coding” functional annotation, with the majority mapping to “introns” (52.7%, i.e., “within a gene”) or “non-gene” (26.2%, i.e., “outside of gene”) annotations. This suggests the presence of large numbers of biologically important variants within proximal or distal elements acting on genes to regulate their expression, where high-throughput functional assays are well positioned to systematically test this set of variants for function (*49*).

Our analysis expands on previous, large-scale fine-mapping experiments (*34, 50, 51*) aimed at determining the candidate causal variant(s), substantially increasing the number of fine-mapped traits and identified signals, particularly for the AFR and AMR populations. The increased representation of diverse participants in our genetic studies also facilitated improved precision of fine-mapping between groups. Notably, the AFR population yielded the most precise approximate credible sets of our three well-powered populations, followed by the EUR and the AMR groups. This finding was anticipated because haplotype blocks are smaller in populations that are genetically similar to African reference populations (*52, 53*). However, the increased precision of the AFR credible set sizes observed in the paired Wilcoxon test did not translate to anticipated differences in the median credible set sizes for signals mapped in the AFR and EUR populations; the median difference was zero. This lack of difference is likely due to the larger sample size in the EUR population increasing statistical power. Thus, we expect that the inclusion of increasing numbers of non-EUR individuals should continue to improve the precision of signal fine-mapping efforts.

The population diversity of MVP enabled a large-scale comparison of the similarities and differences between variants and traits across populations. As anticipated, fine-mapping results identified overwhelmingly more similarities than differences in the genetic associations between groups. The differences observed were largely attributable to variations in allele frequency or the presence of genetic variants in one population that were monomorphic in other populations. This study highlighted a few population-specific variants that may contribute to long-appreciated differences in trait biology. The novel association between the intergenic variant rs7338263 informs longstanding observations of lower WBC and neutrophil counts in individuals genetically similar to the AFR compared to EUR reference populations (*54*), adding to previously identified candidate loci explaining this difference (*29, 55*). The novel signal in the AFR population for rs35965584 with gout in *ABCG2* may represent an independent signal for gout risk, in addition to the known AFR-specific variant rs2231142 in *ABCG2* (*46*). As many risk factors are associated with gout, whether this novel signal contributes to the observed higher prevalence of gout in self-reported Black compared to self-reported White populations requires further study (*56*). Similar trends were seen when considering genome-wide architecture. Genetic heritability of individual traits and genetic correlation between trait pairs supports similar, rather than divergent, cross-population architecture.

Among the most common variants mapped with high precision, there was minimal evidence of heterogeneity in effect estimates. The *APOE* locus was a notable exception, where we observed an association between the high-confidence fine-mapped signal rs429358 and increased risk for dementia across all four populations examined. However, the risk was 30% lower in the AFR compared to EUR groups, consistent with prior studies that observed differential risk between *APOE* alleles and dementia in non-EUR compared to EUR populations (*48, 57*).

Our work must be interpreted within the context of its limitations. To efficiently conduct large-scale GWAS analyses across the phenome, we used an automated approach for phenotyping. This approach involved using Phecodes for collating clinical diagnosis codes and while efficient, could be more precise for most phenotypes. Our regression models also had to be standardized, accounting only for age, sex, and principal components, and performing inverse-normal transformations to quantitative traits prior to analysis. We also encountered challenges in deploying the fine-mapping method at this scale. In particular, the LD matrices used did not ideally synchronize with the SAIGE methodology due to our reliance on hard-called genotypes and not accounting for covariates. This could lead to minor LD mismatches, which may influence sensitive loci, resulting in inaccuracies or spurious results in the fine-mapping stage. Our method of defining loci has potential pitfalls as well. It is based on meta-analyzed data, not considering whether the population-specific GWAS peak was present. Consequently, we fine-mapped some regions with insignificant signals, especially within the EAS population, the smallest population group for which we had limited power. Additionally, we double-counted 115 population-specific signals that overlap between two adjacent loci. Our approach may have also overlooked population-specific peaks eclipsed in the meta-analysis and certain loci too vast to be completely mapped under our scheme. Our conservative approach, adhering to a minimum threshold of significance and purity for signals to maintain precision (positive predictive value), could result in missing true signals. Similarly, our preference for precision over recall (sensitivity) meant we limited the fine-mapping to a maximum of five signals per locus. This approach can lead to an underestimation of the number of signals at certain highly significant loci. Future research may consider these constraints and propose alternative approaches to further enhance the validity and comprehensiveness of the results.

Diversity is a critical feature in advancing precision medicine important in genomic studies because it helps to ensure that the results are generalizable across different populations. Despite large population biobank efforts such as UK Biobank, FinnGen, and Biobank Japan, the limited diversity in the published GWAS literature is quite evident. In recent years, data from more diverse biobanks are becoming become available, such as the All of Us Study Research Program, America Latino Research Biobank, and Human Hereditary and Health in Africa (H3Africa), as well as hospital and institutional biobanks (*8, 10, 58*). Since its inception, the MVP has sought to include a diverse population representative of the diverse United States Veteran population. As of this writing, MVP has enrolled the largest single population of participants genetically similar to African reference populations among current biobanks, expanding our power to study the genetic underpinning of complex traits in a more equitable manner. Our phenome-wide GWAS of health and disease traits among participants in the VA MVP provides a comprehensive atlas of genetic associations across diverse populations. Our overall results highlight the increased power of discovery that the inclusion of individuals from diverse populations brings to the understanding of the genetics of complex health and disease traits.

## Data Availability

The complete summary statistics for all GWAS will soon be accessible to the public. These stats are currently being processed for transfer to dbGAP, and the study's accession number is phs002453.

## Acknowledgments

We thank the Million Veteran Program, Office of Research and Development, and Veterans Health Administration for supporting this work. A complete acknowledgment of contribution to MVP is provided in **Supplementary Note**. We would like to sincerely thank Dr. Thomas Zacharia for providing access to the supercomputers at the Oak Ridge National Laboratory Leadership Computing Facility and Dr. Dimitri Kusenov, the previous DOE Headquarters lead for the VA-DOE partnership, for his invaluable guidance and support. Their contributions have been instrumental in the successful completion of this study. We want to thank NHGRI’s dbGAP team – particularly Mike Feolo, Neha Gupta, Jack Wang, and Anne Sturcke for all their hard work enabling the public release of this large data resource; Gao Wang and Yuxin Zou for their help deriving the formula for residual associations used to tune the parameters for fine-mapping; Colleen Morse for her assistance with visualization. Last but not least, we thank former staff members, and volunteers, who have contributed to MVP and, most of all, MVP participants for their service and their continued contributions to our nation through participation in this study. This publication does not represent the views of the Department of Veteran Affairs or the United States Government.

## Author contributions

### Conceptualization

EB, RR, GT, PST, CJO, SM, KC, MJG, RKM, SD, KPL; **Methodology**: AV, JEH, AR, MC, ML, YLH, YK, DAH, VAP, IG, RT, DCP, RS, MM, XW, DRD, PD, TNN, YS, YVS, SP, AB, WZ, TC, BFV, RKM, SD, KPL; **Investigation**: AV, JEH, AR, MC, ML, TA, CAB, LG, RJC, RC, SD, JG, AH, SKI, JJ, RK, HK, DL, SWL, VCM, CO, JDD, RP, PR, SV, GV, AJ, CJO, AB, TC, BFV, KC, MJG, RKM, SD, KPL; **Visualization**: AV, JEH, AR, MC, LG, VAP, JH, TC, BFV, RKM, SD, KPL**; Funding acquisition:** EB, RR, GT, CJO, SM, MJG, RKM; **Project administration:** YLH, HG, FL, LC, IG, RT, JH, LD, SW, JC, SM, JM, JPC, KC, RKM, SD, KPL; **Supervision:** EB, RR, GT, PST, CJO, TC, BFV, KC, MJG, RKM, SD, KPL; **Writing – original draft:** AV, JEH, AR, MC, ML, BFV, SD, KPL; **Writing – review & editing:** AV, JEH, AR, MC, ML, YLH, YK, DAH, LG, VAP, HG, FL, LC, IG, RT, JH, LD, SW, JC, DCP, RS, MM, XW, DRD, PD, YS, TNN, TA, CAB, RJC, RC, SD, JG, AH, SKI, JJ, RK, HK, DL, SWL, VCM, CO, JDD, SG, RP, PR, YVS, SV, GV, AJ, EB, RR, GT, SP, PST, CJO, SM, JM, JPC, AB, WZ, TC, BFV, KC, MJG, RKM, SD, KPL.

## Data and materials availability

Full summary statistics of all the GWAS are publicly available and can be requested through dbGAP. The dbGAP accession number is phs002453.

## Supplementary Materials

All the supplementary tables and figures can be downloaded from https://tinyurl.com/mww86dn8

## Materials and Methods

### Million Veteran Program study cohort

The VA Million Veteran Program (MVP) is a national cohort launched in 2011 to determine the contributions of genetics, lifestyle, and military exposures to health and disease among US Veterans (*16*). Blood biospecimens were collected for DNA isolation and genotyping. The biorepository was linked with the VA EHR, which includes diagnosis codes (International Classification of Diseases ninth revision [ICD-9] and tenth revision [ICD-10]), laboratory measures, and detailed survey questionnaires collected at the time of enrollment for all veterans and followed in the healthcare system until September 2020.

### Genotyping and imputation

Genotyping and imputation methods for the MVP were described previously (*19*). In brief, the single nucleotide polymorphism (SNP) data in the MVP cohort were generated using a custom ThermoFisher Axiom MVP 1.0 genotyping platform, and imputation was performed to a hybrid reference panel comprised of the African Genome Resources panel (*59*) and 1000 Genomes Project (p3v5) (*60*). Following imputation, variant level quality control (QC) was performed, and genetic variants with 1) imputation quality < 0.3, 2) minor allele count (MAC) < 20, 3) call rate < 97.5% for common variants (minor allele frequency [MAF] > 1%), and 4) call rate < 99% for rare variants (MAF < 1%) were excluded. Additionally, variants were also excluded if they deviated > 10% from their expected allele frequency based on reference data from the 1000 Genomes Project.

### Population group assignment

We used the reference dataset from the 1000 Genomes Project for genetically inferred population estimation with the smartpca module in the EIGENSOFT (*61*) package to project the principal component (PC) loadings from the group of unrelated individuals from the 1000 Genomes Project reference dataset. We merged the 1000 Genomes dataset with the MVP dataset before running smartpca to project the principal component analysis (PCA) loadings from the reference dataset (**fig. S13**). To define the genetically similar population, we trained a random forest classifier using cross-population meta-data based on the top 10 PCs from the reference training data. We used a random forest classifier on the predicted MVP PCA data to assign individuals to one of five 1000 Genomes superpopulation groups. Individuals with random forest probability were over 50% for a population were assigned to that population. Those who could not be assigned to a population were excluded from the study.

### Phenotype data

Clinical outcomes from EHR were defined by Phecodes (*62*), which are curated groupings of ICD codes. Each Phecode represents ICD codes grouped into clinically relevant phenotypes for clinical studies. Case status for binary Phecodes were defined as participants with ≥ 2 instances of the corresponding Phecode-mapped ICD-9 CM or ICD-10 CM codes in the EHR, while control status was the presence of zero instances of mapped ICD codes. Based on our previous studies of ICD EHR data, populations where phenotypes comprise <200 cases or controls are most likely to result in spurious results; this threshold was applied to population-specific analyses. Additionally, the study excluded certain conditions considered protected at the VA, such as sickle cell anemia and HIV status, and their data cannot be reported broadly. Laboratory measures for each participant were summarized as mean, minimum, and maximum across all visits. We filtered values greater than six standard deviations from the mean to remove outliers. Only laboratory traits with more than 1,000 individuals within each population group were included in the analyses. Sixty-nine laboratory traits were included and normalized using rank-based inverse-normal transformation. Survey outcomes were collected from the VA baseline and lifestyle surveys administered at MVP enrollment (*63*) (**Supplementary Note**).

### Genetic association analyses

Within each genetically-inferred population group, we performed a GWAS for each trait of interest to determine the association with each imputed DNA variant. We used the generalized linear mixed model framework to account for participant relatedness and unbalanced case-control ratios with a GPU-optimized version of the SAIGE package implemented on the US Department of Energy’s Summit supercomputer. Directly genotyped variants were used for step 1 of SAIGE. Imputed genetic dosages were used for step 2 of SAIGE. Only variants with an imputation quality > 0.3 and MAC > 20 within the relevant genetically-inferred population groups were included in the GWAS. Analyses were adjusted for age, sex, and ten population-specific genetic PCs.

### Post-GWAS QC

GWAS results were filtered using a custom R script based on EasyQC (*64*). Quality control was implemented to remove variants with missing values for major summary statistics (effect size, standard error, etc.) or with unreasonable values (*P*-values or allele frequencies > 1 or < 0). Variants that were monomorphic, poorly imputed (r^2^ < 0.3), or very rare (MAF < 0.0001) in only the subset of individuals included in the GWAS were also removed. The post-GWAS QC, and subsequent meta-analysis and QC were completed using OLCF’s Andes supercomputing cluster.

### Meta-analysis

Multi-population meta-analysis was performed using the fixed-effect, inverse-variance weighted method implemented in GWAMA (*22*). Meta-analysis results were subjected to the same QC procedures as the GWAS results. Imputation quality filters were not implemented; however, an additional filter excluded variants specific to only one population group.

### Determination of the number of independent traits tested and the genome-wide significance threshold

We calculated Pearson pairwise correlations for the phenotype residuals derived during step 1 of SAIGE to determine the number of independent traits tested. Calculations were made for each population separately and only for the meta-analyzed traits (i.e., passed phenotype QC in more than one population). PCs were calculated from the correlations as well as the variance explained by each of the PCs. The number of independent traits for each population was determined by the number of PCs required to explain a variance of 0.99. For the meta-analysis, population-specific residuals were combined before calculating PCs. This approach resulted in 1,083 independent traits. The population-specific and meta-analysis *P*-value significance thresholds were then determined by dividing the traditional genome-wide threshold of 5 x 10^-8^ by the number of independent traits: 5 × 10^-8^ / 1083, resulting in our study-wide significance of 4.6 × 10^-11^.

### Derivation of independent signals and lead SNPs

For each full GWAS summary statistics dataset, we used PLINK for clumping and thresholding (*65*). A cross-population reference panel was created by selecting 5,000 individuals randomly from each population group. We estimated pairwise correlation within each haplotype block to report independent signals selected those with the smallest *P*-values and counted the number of independent SNPs. The first clumping round grouped SNPs significant at a study-wide *P*-value threshold (i.e., *P* < 4.6 × 10^-11^) and independent at r^2^ < 0.6. This clumping function reported significant, independent SNPs. The second clumping of significant independent SNPs at r^2^ < 0.1 reported the lead SNPs. For both steps, we used a 500-kb distance to compute pairwise LD.

### Defining loci for fine mapping

We tiled the genome into adjoining, non-overlapping 250kb segments and, for each phenotype, we identified all segments that contained one or more variants with a meta-analyzed *P* < 5 × 10^-8^. Due to its long-range LD complexities, we excluded the major histocompatibility complex (MHC) (chr6: 25–36 Mb) from the tiling. Next, within each phenotype, we joined all adjacent significant segments into loci and padded the loci with 250kb on both sides. When significant loci overlapped telomere ends, the telomere-side boundaries of the loci were trimmed to coincide with the chromosomal boundaries. Significant loci overlapping the boundaries of the MHC were similarly trimmed. Finally, we mapped only the loci containing at least one variant significant at the study-wide threshold, *P* = 4.6 × 10^-11^. These parameters for defining loci differed from those used to identify lead variants due to the exclusion of the MHC and the retaining of loci that contained significant short insertions or deletions but not SNPs. Insertions and deletions were later removed from the variants used for fine-mapping to avoid strand-flip issues though the loci were still mapped. Twenty-one traits had no lead SNPs outside the MHC, and 4 had no significant variants besides insertions and deletions.

### Fine-mapping

We statistically fine-mapped the significant phenotype-locus pairs using the Sum of the Single Effects framework (SuSiE) (*32, 65*) with the population-specific summary statistics and computed LD matrices. The LD matrices were matched to each population group and trait to eliminate any potential artifacts that could arise from an LD mismatch between the meta-analyzed GWAS outcomes and the reference panel. We carried out this process independently for each population group, which encompassed AFR, AMR, EAS, and EUR ancestries (**Supplementary Note**). We allowed up to five signals per locus and ran SuSiE through the coloc (*66*) R package with the z-scores as inputs, the “estimate_residual_variance” flag set to true, and the default uniform prior probability of causality. We calculated 95% credible sets for each identified signal representing the fewest number of variants whose posterior inclusion probabilities (PIP) for the signal summed to ≥ 0.95. We discarded credible sets in which the variants had an absolute minimum correlation < 0.1 and/or a minimum *P* > 5 × 10^-8^ for the population group in which the signal was mapped.

We set the maximum number of signals per locus to five instead of the default setting of ten because, during testing, we observed multiple instances of loci with suspicious credible sets at the higher setting. We tested by calculating the residual association for each signal after removing the effects of the other signals according to the following equation:

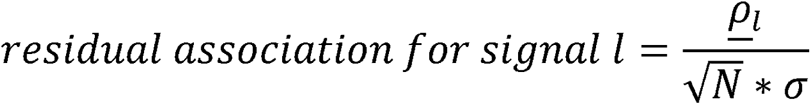

Here <u>ρ_*l*_</u> corresponds to the expected residuals from the SuSiE algorithm after disregarding the effect of the *l*-th signal (see line 4 of the algorithm for Iterative Bayesian stepwise selection using sufficient or summary statistics (*32*)), N is the sample size, and a is the standard deviation for the phenotype under the SuSiE model. Under the null model, the residual association ∼ *N* (0,1).

The suspicious credible sets were non-primary signals at loci for phenotypes with suspected highly polygenic architecture including, most notably, height-related traits. These signals generally manifested as single-variant credible sets whose signal-level residual associations appeared as strong outliers in what would have otherwise been loci with the null residual association for the given signals. Reducing the number of signals mapped per locus from ten to five reduced the frequency of these suspicious credible sets.

### Merging signals across population groups

To identify fine-mapped signals in multiple populations, we merged the retained 95% credible sets using an approach first reported by Kanai *et al.* (*34*). This method involves calculating the PIP-weighted Jaccard similarity indices between all pairs of signals identified for each unique phenotype-locus pair. For each pair of signals, we computed the similarity index as Σ_i_ min(x_i_,y_i_) / Σ_i_ max(x_i_, y_i_), where x_i_ and y_i_ are PIP values for the two signals for each variant *i* that was retained in both populations after making the LD reference panels. We converted each Jaccard similarity index into a distance by taking one minus the similarity index. Then we used the distances to hierarchically cluster the signals using the complete linkage method. We cut the resulting dendrogram tree at the height of 0.9, thereby merging any two credible sets with PIP-weighted Jaccard indices above 0.1 into a single credible set. For any analyses that examined the number of variants in a merged credible set, we defined the variants in each merged approximate credible set as the union of those comprising the component sets.

### Computational environment and constraints

We defined loci, constructed LD matrices, fine-mapping, and signal merging on OLCF’s supercomputing infrastructure. LD matrix construction and fine mapping were parallelized across unique combinations of population, phenotype, and locus, and each combination was given 48 hours of wall time and 1 TB of RAM to complete each step. Any analyses that could not be completed within those constraints or that resulted in an error due to SuSiE estimating a negative residual variance were excluded (**table S8**). All phenotype-locus pairs that appeared in at least one population were put through the signal margining process for the completed populations, and any of their merged signals that passed the absolute minimum correlation and significance thresholds were retained in the fine-mapping results.

### Suggestive associations and power calculations

To investigate the cause of a large number of signals mapped in a single population group, we looked for signals that had suggestive evidence of an association or lacked the power to detect a suggestive association in one or more unmapped populations. Signals were considered to show suggestive evidence of association in an unmapped population if any variants in the merged approximate credible set had directionally consistent effects between an unmapped and mapped population and population-specific *P* < 0.001 in the unmapped population. We also calculated the approximate power to identify associations in the unmapped populations using a two-step approach based on the t-distribution. In the first step, we calculated the critical effect size (absolute beta) for each variant within each unmapped population above which we would detect a suggestive association, given the standard errors and sample sizes for the unmapped population groups. In the second step, we assessed the proportion of an identically shaped t-distribution centered at the “true” effect size for each variant that lay above the critical value or below its negative opposite. For the “true” effect size of a given variant, we assumed the smallest absolute beta across the populations in which the variant was mapped in the credible set. If the proportion outside the critical values was < 80% in any of the unmapped populations and the signal did not show suggestive association, we considered that the signal lacked power.

### Effective sample size calculations

To jointly examine the relationship between sample size and the number of fine-mapped signals for quantitative and binary traits, we calculated effective sample sizes for binary traits as the harmonic mean of the cross-population case and control counts (*67*) for the trait. For quantitative traits, we similarly used the cross-population sample sizes.

### Functional annotations and coding enrichment

We used the Variant Effect Predictor (VEP) (*68*) to determine the most severe functional consequence of each associated and fine-mapped variant. The 39 annotations assigned by VEP were grouped into ten larger categories, including three comprising coding variants: splice/start/stop gain/loss, missense, and synonymous (**table S12**). For each variant annotated as non-coding, we used RegulomeDB v2.2 (*35*) to assign a probability score and category indicating the variant’s probability of being functional. To assess enrichment in coding variation and higher RegulomeDB scores across different categories of variants, we respectively used Fisher’s exact and Wilcoxon rank sum tests.

### Derived and ancestral allele identification for high-confidence, fine-mapped variants

We determined the identities of the ancestral and derived alleles for all variants that were fine-mapped with high confidence in any population (PIP > 0.95) by referencing the parsimony-derived ancestral allele identities in the 1000 Genomes (1000G) Phase III variant call format files (*60*). Given our imputation scheme, 6,150 of 6,318 (97.3%) of the high-confidence variants were included in the 1000G callsets. Of the included variants, only those with consistently annotated ancestral alleles across the inferred orangutan-chimp-human progenitor, inferred chimp-human progenitor, and chimp lineages were retained for analyses involving ancestral/derived alleles. These remaining variants comprise 12,613 high-confidence SNP-phenotype pairs.

### Defining known and novel associations

We compared the lead and fine-mapped variants and their proxies (with an r2 > 0.1) to determine their previously reported association in both the GWAS Catalog and Open Target Genetics database. These databases employ Experimental Factor Ontology (EFO) terms as the principal vocabulary for standardizing traits and phenotypes, prompting us to initially map all traits analyzed in our study to EFO terms. We implemented semi-automated processes to map specific trait descriptions to EFO terms to bridge the gap between disparate data sources. This process involves processing various mapping files and EFO terms from the GWAS catalog and Open Target Genetics, which provide Phecode, labs, and vitals to EFO term mappings. Initially, we map these Phecodes to their corresponding EFO terms using pre-existing mappings. For traits not categorized as Phecodes, we employed a “text descriptive fuzzy mapping” technique to assign these remaining traits to EFO terms. Subsequently, we carried out a manual review of trait to EFO term mappings since there were instances where different EFO terms were used for the same term by NHGRI EMBL GWAS (*24*) and Open Target Genetics catalogs (*69*). Upon a thorough review, the final table was composed, including traits and their corresponding EFO terms, ready to cross-reference with the GWAS catalog and Open Target Genetics.

For each trait, we identified the tag SNPs associated with each lead SNP (r^2^ > 0.1 and within a 500kb window). We then proceeded to cross-reference the lead SNP and the tag SNP in the GWAS catalog and Open Target Genetics to determine if they were associated with the trait of interest. We categorized the novelty of each association into one of three groups: a) Known Association: when either the lead SNP or tag SNPs are associated with the same trait as recorded in the GWAS or Open Target Genetics catalogs; b) Novel Association with Known Signal: when the lead SNP or tag SNPs are not associated with the same trait but do have associations with other traits; c) Novel Signal: when the lead SNP or tag SNPs are not associated with any trait listed in the GWAS catalog or Open Target Genetics.

### Cross-population comparisons of credible set sizes

We compared the number of variants in the population-specific credible sets for signals successfully merged across more than one population (excluding the EAS population, as it lacked sufficient power). We used a paired Wilcoxon sign-rank test to identify significant differences in the credible set sizes for signals mapped in AFR/AMR, AFR/EUR, and AMR/EUR populations.

### Identifying variant-trait associations specific to the non-EUR populations

We focused on characterizing novel SNPs in the non-EUR population fine-mapped to a trait by identifying variants mapped with high confidence (PIP>0.95) in the non-EUR populations and either unmapped or mapped with low confidence in the EUR population (**table S11**). We further restricted to coding variants as defined by VEP and with a MAF>0.05 in each of the non-EUR populations in the main results.

### Estimating heterogeneous effects

Screening for heterogeneous effects across populations was performed on variants fine-mapped to a trait, specifically for Phecode only, from any population with a MAF>0.05. The quantitative traits were not tested for heterogeneity due to the potential discrepancy in their scales across different population groups. The variant-trait pair must have been fine-mapped with high confidence (PIP>0.95) in both populations being compared (AFR vs. EUR, AMR vs. EUR; the sample size for EAS was underpowered for the multiple testing among ultra-high-dimensional hypotheses) with MAF>0.05. To adequately control for the false discovery rate (FDR), the heterogeneity analysis performs an adaptive multiple testing procedure on the heterogeneity effect (*69*) (**Supplementary Note**) on the full set of variant-phenotype pairs. The adaptive heterogeneity multiple testing procedure improves power by reweighting the heterogeneity test statistics according to the level of evidence for the presence of an overall mean effect. In the broad heterogeneity screen, we examined all fine-mapped variants with MAF>0.05 in both populations being compared, i.e., AFR vs. EUR and AMR vs. EUR, and required overlapping credible sets. This allowed us to potentially detect variants with effects only present in the population or with small effect sizes that may be underpowered for AFR or AMR populations. The primary heterogeneity analysis further required that the variant be amino acid changing with PIP>0.95 in both populations and that the fine-mapping result showed the same variant-phenotype pair in both populations.

### Selection of pleiotropic associations

To assess pleiotropy for SNPs with more than one trait association and PIP > 0.95, we needed to determine which traits were correlated. For each SNP, the following procedure was used to determine the number of independent traits. First, a trait (trait #1) was selected for comparison with all others based on the largest PIP value (0.95–1). If there was a tie, we selected trait #1 as (a) the trait with the most significant meta-analysis *P*-value, (b) the most significant population-specific P-value starting with the largest population size (EUR, AFR, AMR, then EAS), or (c) the trait with the largest absolute meta-analysis effect estimate. Next, all associated traits with an absolute phenotypic correlation coefficient >0.2 with trait #1 were flagged as correlated. We iterated over these steps until only uncorrelated traits remained. (**fig. S12**).

**Supplementary Note**

**Figs. S1 to S14**

**Tables S1 to S19**

## References and Notes

1. M. C. Mills, C. Rahal, The GWAS Diversity Monitor tracks diversity by disease in real time. Nat. Genet. 52, 242–243 (2020).

2. A. R. Martin, M. Kanai, Y. Kamatani, Y. Okada, B. M. Neale, M. J. Daly, Clinical use of current polygenic risk scores may exacerbate health disparities. Nat. Genet. 51, 584–591 (2019).

3. S. L. Pulit, B. F. Voight, P. I. W. de Bakker, Multiethnic genetic association studies improve power for locus discovery. PloS One. 5, e12600 (2010).

4. G. Genovese, D. J. Friedman, M. D. Ross, L. Lecordier, P. Uzureau, B. I. Freedman, D. W. Bowden, C. D. Langefeld, T. K. Oleksyk, A. L. Uscinski Knob, A. J. Bernhardy, P. J. Hicks, G. W. Nelson, B. Vanhollebeke, C. A. Winkler, J. B. Kopp, E. Pays, M. R. Pollak, Association of trypanolytic ApoL1 variants with kidney disease in African Americans. Science. 329, 841–845 (2010).

5. E. G. Cohn, G. E. Henderson, P. S. Appelbaum, Distributive justice, diversity, and inclusion in precision medicine: what will success look like? Genet. Med. 19, 157–159 (2017).

6. B. M. Mapes, C. S. Foster, S. V. Kusnoor, M. I. Epelbaum, M. AuYoung, G. Jenkins, M. Lopez-Class, D. Richardson-Heron, A. Elmi, K. Surkan, R. M. Cronin, C. H. Wilkins, E. J. Pérez-Stable, E. Dishman, J. C. Denny, J. L. Rutter, the All of Us Research Program, Diversity and inclusion for the All of Us research program: A scoping review. PLOS ONE. 15, e0234962 (2020).

7. B. N. Wolford, C. J. Willer, I. Surakka, Electronic health records: the next wave of complex disease genetics. Hum. Mol. Genet. 27, R14–R21 (2018).

8. A. Verma, S. M. Damrauer, N. Naseer, J. Weaver, C. M. Kripke, L. Guare, G. Sirugo, R. L. Kember, T. G. Drivas, S. M. Dudek, Y. Bradford, A. Lucas, R. Judy, S. S. Verma, E. Meagher, K. L. Nathanson, M. Feldman, M. D. Ritchie, D. J. Rader, For The Penn Medicine BioBank, The Penn Medicine BioBank: Towards a Genomics-Enabled Learning Healthcare System to Accelerate Precision Medicine in a Diverse Population. J. Pers. Med. 12, 1974 (2022).

9. C. Sudlow, J. Gallacher, N. Allen, V. Beral, P. Burton, J. Danesh, P. Downey, P. Elliott, J. Green, M. Landray, B. Liu, P. Matthews, G. Ong, J. Pell, A. Silman, A. Young, T. Sprosen, T. Peakman, R. Collins, UK biobank: an open access resource for identifying the causes of a wide range of complex diseases of middle and old age. PLoS Med. 12, e1001779 (2015).

10. M. Zawistowski, L. G. Fritsche, A. Pandit, B. Vanderwerff, S. Patil, E. M. Schmidt, P. VandeHaar, C. J. Willer, C. M. Brummett, S. Kheterpal, X. Zhou, M. Boehnke, G. R. Abecasis, S. Zöllner, The Michigan Genomics Initiative: A biobank linking genotypes and electronic clinical records in Michigan Medicine patients. Cell Genomics. 3, 100257 (2023).

11. M. I. Kurki, J. Karjalainen, P. Palta, T. P. Sipilä, K. Kristiansson, K. M. Donner, M. P. Reeve, H. Laivuori, M. Aavikko, M. A. Kaunisto, A. Loukola, E. Lahtela, H. Mattsson, P. Laiho, P. Della Briotta Parolo, A. A. Lehisto, M. Kanai, N. Mars, J. Rämö, T. Kiiskinen, H. O. Heyne, K. Veerapen, S. Rüeger, S. Lemmelä, W. Zhou, S. Ruotsalainen, K. Pärn, T. Hiekkalinna, S. Koskelainen, T. Paajanen, V. Llorens, J. Gracia-Tabuenca, H. Siirtola, K. Reis, A. G. Elnahas, B. Sun, C. N. Foley, K. Aalto-Setälä, K. Alasoo, M. Arvas, K. Auro, S. Biswas, A. Bizaki-Vallaskangas, O. Carpen, C.-Y. Chen, O. A. Dada, Z. Ding, M. G. Ehm, K. Eklund, M. Färkkilä, H. Finucane, A. Ganna, A. Ghazal, R. R. Graham, E. M. Green, A. Hakanen, M. Hautalahti, Å. K. Hedman, M. Hiltunen, R. Hinttala, I. Hovatta, X. Hu, A. Huertas-Vazquez, L. Huilaja, J. Hunkapiller, H. Jacob, J.-N. Jensen, H. Joensuu, S. John, V. Julkunen, M. Jung, J. Junttila, K. Kaarniranta, M. Kähönen, R. Kajanne, L. Kallio, R. Kälviäinen, J. Kaprio, FinnGen, N. Kerimov, J. Kettunen, E. Kilpeläinen, T. Kilpi, K. Klinger, V-M. Kosma, T. Kuopio, V. Kurra, T. Laisk, J. Laukkanen, N. Lawless, A. Liu, S. Longerich, R. Mägi, J. Mäkelä, A. Mäkitie, A. Malarstig, A. Mannermaa, J. Maranville, A. Matakidou, T. Meretoja, S. V. Mozaffari, M. E. K. Niemi, M. Niemi, T. Niiranen, C. J. ÓDonnell, M. Obeidat, G. Okafo, H. M. Ollila, A. Palomäki, T. Palotie, J. Partanen, D. S. Paul, M. Pelkonen, R. K. Pendergrass, S. Petrovski, A. Pitkäranta, A. Platt, D. Pulford, E. Punkka, P. Pussinen, N. Raghavan, F. Rahimov, D. Rajpal, N. A. Renaud, B. Riley-Gillis, R. Rodosthenous, E. Saarentaus, A. Salminen, E. Salminen, V. Salomaa, J. Schleutker, R. Serpi, H. Shen, R. Siegel, K. Silander, S. Siltanen, S. Soini, H. Soininen, J. H. Sul, I. Tachmazidou, K. Tasanen, P. Tienari, S. Toppila-Salmi, T. Tukiainen, T. Tuomi, J. A. Turunen, J. C. Ulirsch, F. Vaura, P. Virolainen, J. Waring, D. Waterworth, R. Yang, M. Nelis, A. Reigo, A. Metspalu, L. Milani, T. Esko, C. Fox, A. S. Havulinna, M. Perola, S. Ripatti, A. Jalanko, T. Laitinen, T. P. Mäkelä, R. Plenge, M. McCarthy, H. Runz, M. J. Daly, A. Palotie, FinnGen provides genetic insights from a well-phenotyped isolated population. Nature. 613, 508–518 (2023).

12. O. Gottesman, H. Kuivaniemi, G. Tromp, W. A. Faucett, R. Li, T. A. Manolio, S. C. Sanderson, J. Kannry, R. Zinberg, M. A. Basford, M. Brilliant, D. J. Carey, R. L. Chisholm, C. G. Chute, J. J. Connolly, D. Crosslin, J. C. Denny, C. J. Gallego, J. L. Haines, H. Hakonarson, J. Harley, G. P. Jarvik, I. Kohane, I. J. Kullo, E. B. Larson, C. McCarty, M. D. Ritchie, D. M. Roden, M. E. Smith, E. P. Böttinger, M. S. Williams, The Electronic Medical Records and Genomics (eMERGE) Network: past, present, and future. Genet. Med. 15, 761–771 (2013).

13. H. Carress, D. J. Lawson, E. Elhaik, Population genetic considerations for using biobanks as international resources in the pandemic era and beyond. BMC Genomics. 22, 351 (2021).

14. W. Zhou, M. Kanai, K.-H. H. Wu, H. Rasheed, K. Tsuo, J. B. Hirbo, Y. Wang, A. Bhattacharya, H. Zhao, S. Namba, I. Surakka, B. N. Wolford, V. Lo Faro, E. A. Lopera-Maya, K. Läll, M.-J. Favé, J. J. Partanen, S. B. Chapman, J. Karjalainen, M. Kurki, M. Maasha, B. M. Brumpton, S. Chavan, T.-T. Chen, M. Daya, Y. Ding, Y.-C. A. Feng, L. A. Guare, C. R. Gignoux, S. E. Graham, W. E. Hornsby, N. Ingold, S. I. Ismail, R. Johnson, T. Laisk, K. Lin, J. Lv, I. Y. Millwood, S. Moreno-Grau, K. Nam, P. Palta, A. Pandit, M. H. Preuss, C. Saad, S. Setia-Verma, U. Thorsteinsdottir, J. Uzunovic, A. Verma, M. Zawistowski, X. Zhong, N. Afifi, K. M. Al-Dabhani, A. Al Thani, Y. Bradford, A. Campbell, K. Crooks, G. H. de Bock, S. M. Damrauer, N. J. Douville, S. Finer, L. G. Fritsche, E. Fthenou, G. Gonzalez-Arroyo, C. J. Griffiths, Y. Guo, K. A. Hunt, A. Ioannidis, N. M. Jansonius, T. Konuma, M. T. M. Lee, A. Lopez-Pineda, Y. Matsuda, R. E. Marioni, B. Moatamed, M. A. Nava-Aguilar, K. Numakura, S. Patil, N. Rafaels, A. Richmond, A. Rojas-Muñoz, J. A. Shortt, P. Straub, R. Tao, B. Vanderwerff, M. Vernekar, Y. Veturi, K. C. Barnes, M. Boezen, Z. Chen, C.-Y. Chen, J. Cho, G. D. Smith, H. K. Finucane, L. Franke, E. R. Gamazon, A. Ganna, T. R. Gaunt, T. Ge, H. Huang, J. Huffman, N. Katsanis, J. T. Koskela, C. Lajonchere, M. H. Law, L. Li, C. M. Lindgren, R. J. F. Loos, S. MacGregor, K. Matsuda, C. M. Olsen, D. J. Porteous, J. A. Shavit, H. Snieder, T. Takano, R. C. Trembath, J. M. Vonk, D. C. Whiteman, S. J. Wicks, C. Wijmenga, J. Wright, J. Zheng, X. Zhou, P. Awadalla, M. Boehnke, C. D. Bustamante, N. J. Cox, S. Fatumo, D. H. Geschwind, C. Hayward, K. Hveem, E. E. Kenny, S. Lee, Y.-F. Lin, H. Mbarek, R. Mägi, H. C. Martin, S. E. Medland, Y. Okada, A. V. Palotie, B. Pasaniuc, D. J. Rader, M. D. Ritchie, S. Sanna, J. W. Smoller, K. Stefansson, D. A. van Heel, R. G. Walters, S. Zöllner, A. R. Martin, C. J. Willer, M. J. Daly, B. M. Neale, Global Biobank Meta-analysis Initiative: Powering genetic discovery across human disease. Cell Genomics. 2, 100192 (2022).

15. Z. Chen, J. Chen, R. Collins, Y. Guo, R. Peto, F. Wu, L. Li, China Kadoorie Biobank (CKB) collaborative group, China Kadoorie Biobank of 0.5 million people: survey methods, baseline characteristics and long-term follow-up. Int. J. Epidemiol. 40, 1652–1666 (2011).

16. J. M. Gaziano, J. Concato, M. Brophy, L. Fiore, S. Pyarajan, J. Breeling, S. Whitbourne, J. Deen, C. Shannon, D. Humphries, P. Guarino, M. Aslan, D. Anderson, R. LaFleur, T. Hammond, K. Schaa, J. Moser, G. Huang, S. Muralidhar, R. Przygodzki, T. J. O’Leary, Million Veteran Program: A mega-biobank to study genetic influences on health and disease. J. Clin. Epidemiol. 70, 214–223 (2016).

17. J.-E. Lee, J.-H. Kim, E.-J. Hong, H. S. Yoo, H.-Y. Nam, O. Park, National Biobank of Korea: Quality control Programs of Collected-human Biospecimens. Osong Public Health Res. Perspect. 3, 185–189 (2012).

18. All of Us Research Program Investigators, J. C. Denny, J. L. Rutter, D. B. Goldstein, A. Philippakis, J. W. Smoller, G. Jenkins, E. Dishman, The “All of Us” Research Program. N. Engl. J. Med. 381, 668–676 (2019).

19. H. Hunter-Zinck, Y. Shi, M. Li, B. R. Gorman, S.-G. Ji, N. Sun, T. Webster, A. Liem, P. Hsieh, P. Devineni, P. Karnam, X. Gong, L. Radhakrishnan, J. Schmidt, T. L. Assimes, J. Huang, C. Pan, D. Humphries, M. Brophy, J. Moser, S. Muralidhar, G. D. Huang, R. Przygodzki, J. Concato, J. M. Gaziano, J. Gelernter, C. J. O’Donnell, E. R. Hauser, H. Zhao, T. J. O’Leary, VA Million Veteran Program, P. S. Tsao, S. Pyarajan, Genotyping Array Design and Data Quality Control in the Million Veteran Program. Am. J. Hum. Genet. 106, 535–548 (2020).

20. Committee on the Use of Race, Ethnicity, and Ancestry as Population Descriptors in Genomics Research, Board on Health Sciences Policy, Committee on Population, Health and Medicine Division, Division of Behavioral and Social Sciences and Education, National Academies of Sciences, Engineering, and Medicine, Using Population Descriptors in Genetics and Genomics Research: A New Framework for an Evolving Field (National Academies Press, Washington, D.C., 2023; https://www.nap.edu/catalog/26902).

21. W. Zhou, J. B. Nielsen, L. G. Fritsche, R. Dey, M. E. Gabrielsen, B. N. Wolford, J. LeFaive, P. VandeHaar, S. A. Gagliano, A. Gifford, L. A. Bastarache, W.-Q. Wei, J. C. Denny, M. Lin, K. Hveem, H. M. Kang, G. R. Abecasis, C. J. Willer, S. Lee, Efficiently controlling for case-control imbalance and sample relatedness in large-scale genetic association studies. Nat. Genet. 50, 1335–1341 (2018).

22. R. Mägi, A. P. Morris, GWAMA: software for genome-wide association meta-analysis. BMC Bioinformatics. 11, 288 (2010).

23. F. Zhang, J. R. Lupski, Non-coding genetic variants in human disease: Figure 1. Hum. Mol. Genet. 24, R102–R110 (2015).

24. E. Sollis, A. Mosaku, A. Abid, A. Buniello, M. Cerezo, L. Gil, T. Groza, O. Güneş, P. Hall, J. Hayhurst, A. Ibrahim, Y. Ji, S. John, E. Lewis, J. A. L. MacArthur, A. McMahon, D. Osumi-Sutherland, K. Panoutsopoulou, Z. Pendlington, S. Ramachandran, R. Stefancsik, J. Stewart, P. Whetzel, R. Wilson, L. Hindorff, F. Cunningham, S. A. Lambert, M. Inouye, H. Parkinson, L. W. Harris, The NHGRI-EBI GWAS Catalog: knowledgebase and deposition resource. Nucleic Acids Res. 51, D977–D985 (2023).

25. M. Ghoussaini, E. Mountjoy, M. Carmona, G. Peat, E. M. Schmidt, A. Hercules, L. Fumis, A. Miranda, D. Carvalho-Silva, A. Buniello, T. Burdett, J. Hayhurst, J. Baker, J. Ferrer, A. Gonzalez-Uriarte, S. Jupp, M. A. Karim, G. Koscielny, S. Machlitt-Northen, C. Malangone, Z. M. Pendlington, P. Roncaglia, D. Suveges, D. Wright, O. Vrousgou, E. Papa, H. Parkinson, J. A. L. MacArthur, J. A. Todd, J. C. Barrett, J. Schwartzentruber, D. G. Hulcoop, D. Ochoa, E. M. McDonagh, I. Dunham, Open Targets Genetics: systematic identification of trait-associated genes using large-scale genetics and functional genomics. Nucleic Acids Res. 49, D1311–D1320 (2021).

26. B. F. Darst, P. Wan, X. Sheng, J. T. Bensen, S. A. Ingles, B. A. Rybicki, B. Nemesure, E. M. John, J. H. Fowke, V. L. Stevens, S. I. Berndt, C. D. Huff, S. S. Strom, J. Y. Park, W. Zheng, E. A. Ostrander, P. C. Walsh, S. Srivastava, J. Carpten, T. A. Sellers, K. Yamoah, A. B. Murphy, M. Sanderson, D. C. Crawford, S. M. Gapstur, W. S. Bush, M. C. Aldrich, O. Cussenot, M. Yeager, G. Petrovics, J. Cullen, C. Neslund-Dudas, R. A. Kittles, J. Xu, M. C. Stern, Z. Kote-Jarai, K. Govindasami, A. P. Chokkalingam, L. Multigner, M.-E. Parent, F. Menegaux, G. Cancel-Tassin, A. S. Kibel, E. A. Klein, P. J. Goodman, B. F. Drake, J. J. Hu, P. E. Clark, P. Blanchet, G. Casey, A. J. M. Hennis, A. Lubwama, I. M. Thompson, R. Leach, S. M. Gundell, L. Pooler, L. Xia, J. L. Mohler, E. T. H. Fontham, G. J. Smith, J. A. Taylor, R. A. Eeles, L. Brureau, S. J. Chanock, S. Watya, J. L. Stanford, D. Mandal, W. B. Isaacs, K. Cooney, W. J. Blot, D. V. Conti, C. A. Haiman, A Germline Variant at 8q24 Contributes to Familial Clustering of Prostate Cancer in Men of African Ancestry. Eur. Urol. 78, 316–320 (2020).

27. O. A. Panagiotou, R. C. Travis, D. Campa, S. I. Berndt, S. Lindstrom, P. Kraft, F. R. Schumacher, A. Siddiq, S. I. Papatheodorou, J. L. Stanford, D. Albanes, J. Virtamo, S. J. Weinstein, W. R. Diver, S. M. Gapstur, V. L. Stevens, H. Boeing, H. B. Bueno-de-Mesquita, A. Barricarte Gurrea, R. Kaaks, K.-T. Khaw, V. Krogh, K. Overvad, E. Riboli, D. Trichopoulos, E. Giovannucci, M. Stampfer, C. Haiman, B. Henderson, L. Le Marchand, J. M. Gaziano, D. J. Hunter, S. Koutros, M. Yeager, R. N. Hoover, S. J. Chanock, S. Wacholder, T. J. Key, K. K. Tsilidis, A Genome-wide Pleiotropy Scan for Prostate Cancer Risk. Eur. Urol. 67, 649–657 (2015).

28. F. Chen, R. K. Madduri, A. A. Rodriguez, B. F. Darst, A. Chou, X. Sheng, A. Wang, J. Shen, E. J. Saunders, S. K. Rhie, J. T. Bensen, S. A. Ingles, R. A. Kittles, S. S. Strom, B. A. Rybicki, B. Nemesure, W. B. Isaacs, J. L. Stanford, W. Zheng, M. Sanderson, E. M. John, J. Y. Park, J. Xu, Y. Wang, S. I. Berndt, C. D. Huff, E. D. Yeboah, Y. Tettey, J. Lachance, W. Tang, C. T. Rentsch, K. Cho, B. H. Mcmahon, R. B. Biritwum, A. A. Adjei, E. Tay, A. Truelove, S. Niwa, T. A. Sellers, K. Yamoah, A. B. Murphy, D. C. Crawford, A. V. Patel, W. S. Bush, M. C. Aldrich, O. Cussenot, G. Petrovics, J. Cullen, C. M. Neslund-Dudas, M. C. Stern, Z. Kote-Jarai, K. Govindasami, M. B. Cook, A. P. Chokkalingam, A. W. Hsing, P. J. Goodman, T. J. Hoffmann, B. F. Drake, J. J. Hu, J. M. Keaton, J. N. Hellwege, P. E. Clark, M. Jalloh, S. M. Gueye, L. Niang, O. Ogunbiyi, M. O. Idowu, O. Popoola, A. O. Adebiyi, O. I. Aisuodionoe-Shadrach, H. O. Ajibola, M. A. Jamda, O. P. Oluwole, M. Nwegbu, B. Adusei, S. Mante, A. Darkwa-Abrahams, J. E. Mensah, H. Diop, S. K. Van Den Eeden, P. Blanchet, J. H. Fowke, G. Casey, A. J. Hennis, A. Lubwama, I. M. Thompson, R. Leach, D. F. Easton, M. H. Preuss, R. J. Loos, S. M. Gundell, P. Wan, J. L. Mohler, E. T. Fontham, G. J. Smith, J. A. Taylor, S. Srivastava, R. A. Eeles, J. D. Carpten, A. S. Kibel, L. Multigner, M.-É. Parent, F. Menegaux, G. Cancel-Tassin, E. A. Klein, C. Andrews, T. R. Rebbeck, L. Brureau, S. Ambs, T. L. Edwards, S. Watya, S. J. Chanock, J. S. Witte, W. J. Blot, J. Michael Gaziano, A. C. Justice, D. V. Conti, C. A. Haiman, Evidence of Novel Susceptibility Variants for Prostate Cancer and a Multiancestry Polygenic Risk Score Associated with Aggressive Disease in Men of African Ancestry. Eur. Urol., S0302–2838(23)02561–7 (2023).

29. D. Reich, M. A. Nalls, W. H. L. Kao, E. L. Akylbekova, A. Tandon, N. Patterson, J. Mullikin, W.-C. Hsueh, C.-Y. Cheng, J. Coresh, E. Boerwinkle, M. Li, A. Waliszewska, J. Neubauer, R. Li, T. S. Leak, L. Ekunwe, J. C. Files, C. L. Hardy, J. M. Zmuda, H. A. Taylor, E. Ziv, T. B. Harris, J. G. Wilson, Reduced Neutrophil Count in People of African Descent Is Due To a Regulatory Variant in the Duffy Antigen Receptor for Chemokines Gene. PLoS Genet. 5, e1000360 (2009).

30. A. R. Bentley, J. Divers, D. Shriner, A. P. Doumatey, O. M. Gutiérrez, A. A. Adeyemo, B. I. Freedman, C. N. Rotimi, APOL1 G1 genotype modifies the association between HDLC and kidney function in African Americans. BMC Genomics. 16, 421 (2015).

31. T. P. Joshi, D. Garcia, F. Gedeon, D. Hinson, E. Strouphauer, F. Okundia, J. Tschen, Epidemiology of alopecia areata in the Hispanic/Latinx community: A cross-sectional analysis of the All of Us database. J. Am. Acad. Dermatol., S0190962223003717 (2023).

32. G. Wang, A. Sarkar, P. Carbonetto, M. Stephens, A Simple New Approach to Variable Selection in Regression, with Application to Genetic Fine Mapping. J. R. Stat. Soc. Ser. B Stat. Methodol. 82, 1273–1300 (2020).

33. Y. Zou, P. Carbonetto, G. Wang, M. Stephens, Fine-mapping from summary data with the “Sum of Single Effects” model. PLOS Genet. 18, e1010299 (2022).

34. M. Kanai, J. C. Ulirsch, J. Karjalainen, M. Kurki, K. J. Karczewski, E. Fauman, Q. S. Wang, H. Jacobs, F. Aguet, K. G. Ardlie, N. Kerimov, K. Alasoo, C. Benner, K. Ishigaki, S. Sakaue, S. Reilly, The BioBank Japan Project, FinnGen, Y. Kamatani, K. Matsuda, A. Palotie, B. M. Neale, R. Tewhey, P. C. Sabeti, Y. Okada, M. J. Daly, H. K. Finucane, “Insights from complex trait fine-mapping across diverse populations” (preprint, Genetic and Genomic Medicine, 2021), doi:10.1101/2021.09.03.21262975.

35. S. Dong, N. Zhao, E. Spragins, M. S. Kagda, M. Li, P. Assis, O. Jolanki, Y. Luo, J. M. Cherry, A. P. Boyle, B. C. Hitz, Annotating and prioritizing human non-coding variants with RegulomeDB v.2. Nat. Genet. 55, 724–726 (2023).

36. J.-H. Park, M. H. Gail, C. R. Weinberg, R. J. Carroll, C. C. Chung, Z. Wang, S. J. Chanock, J. F. Fraumeni, N. Chatterjee, Distribution of allele frequencies and effect sizes and their interrelationships for common genetic susceptibility variants. Proc. Natl. Acad. Sci. U. S. A. 108, 18026–18031 (2011).

37. A. P. Schoech, D. M. Jordan, P.-R. Loh, S. Gazal, L. J. O’Connor, D. J. Balick, P. F. Palamara, H. K. Finucane, S. R. Sunyaev, A. L. Price, Quantification of frequency-dependent genetic architectures in 25 UK Biobank traits reveals action of negative selection. Nat. Commun. 10, 790 (2019).

38. G. Wang, J. R. Speakman, Analysis of Positive Selection at Single Nucleotide Polymorphisms Associated with Body Mass Index Does Not Support the “Thrifty Gene” Hypothesis. Cell Metab. 24, 531–541 (2016).

39. S. Wilde, A. Timpson, K. Kirsanow, E. Kaiser, M. Kayser, M. Unterländer, N. Hollfelder, I. D. Potekhina, W. Schier, M. G. Thomas, J. Burger, Direct evidence for positive selection of skin, hair, and eye pigmentation in Europeans during the last 5,000 y. Proc. Natl. Acad. Sci. 111, 4832–4837 (2014).

40. R. L. Lamason, M.-A. P. K. Mohideen, J. R. Mest, A. C. Wong, H. L. Norton, M. C. Aros, M. J. Jurynec, X. Mao, V. R. Humphreville, J. E. Humbert, S. Sinha, J. L. Moore, P. Jagadeeswaran, W. Zhao, G. Ning, I. Makalowska, P. M. McKeigue, D. O’Donnell, R. Kittles, E. J. Parra, N. J. Mangini, D. J. Grunwald, M. D. Shriver, V. A. Canfield, K. C. Cheng, SLC24A5, a Putative Cation Exchanger, Affects Pigmentation in Zebrafish and Humans. Science. 310, 1782–1786 (2005).

41. L. Yengo, S. Vedantam, E. Marouli, J. Sidorenko, E. Bartell, S. Sakaue, M. Graff, A. U. Eliasen, Y. Jiang, S. Raghavan, J. Miao, J. D. Arias, S. E. Graham, R. E. Mukamel, C. N. Spracklen, X. Yin, S.-H. Chen, T. Ferreira, H. H. Highland, Y. Ji, T. Karaderi, K. Lin, K. Lüll, D. E. Malden, C. Medina-Gomez, M. Machado, A. Moore, S. Rüeger, X. Sim, S. Vrieze, T. S. Ahluwalia, M. Akiyama, M. A. Allison, M. Alvarez, M. K. Andersen, A. Ani, V. Appadurai, L. Arbeeva, S. Bhaskar, L. F. Bielak, S. Bollepalli, L. L. Bonnycastle, J. Bork-Jensen, J. P. Bradfield, Y. Bradford, P. S. Braund, J. A. Brody, K. S. Burgdorf, B. E. Cade, H. Cai, Q. Cai, A. Campbell, M. Cañadas-Garre, E. Catamo, J.-F. Chai, X. Chai, L.-C. Chang, Y.-C. Chang, C.-H. Chen, A. Chesi, S. H. Choi, R.-H. Chung, M. Cocca, M. P. Concas, C. Couture, G. Cuellar-Partida, R. Danning, E. W. Daw, F. Degenhard, G. E. Delgado, A. Delitala, A. Demirkan, X. Deng, P. Devineni, A. Dietl, M. Dimitriou, L. Dimitrov, R. Dorajoo, A. B. Ekici, J. E. Engmann, Z. Fairhurst-Hunter, A.-E. Farmaki, J. D. Faul, J.-C. Fernandez-Lopez, L. Forer, M. Francescatto, S. Freitag-Wolf, C. Fuchsberger, T. E. Galesloot, Y. Gao, Z. Gao, F. Geller, O. Giannakopoulou, F. Giulianini, A. P. Gjesing, A. Goel, S. D. Gordon, M. Gorski, J. Grove, X. Guo, S. Gustafsson, J. Haessler, T. F. Hansen, A. S. Havulinna, S. J. Haworth, J. He, N. Heard-Costa, P. Hebbar, G. Hindy, Y.-L. A. Ho, E. Hofer, E. Holliday, K. Horn, W. E. Hornsby, J.-J. Hottenga, H. Huang, J. Huang, A. Huerta-Chagoya, J. E. Huffman, Y.-J. Hung, S. Huo, M. Y. Hwang, H. Iha, D. D. Ikeda, M. Isono, A. U. Jackson, S. Jäger, I. E. Jansen, I. Johansson, J. B. Jonas, A. Jonsson, T. Jørgensen, I.-P. Kalafati, M. Kanai, S. Kanoni, L. L. Kårhus, A. Kasturiratne, T. Katsuya, T. Kawaguchi, R. L. Kember, K. A. Kentistou, H.-N. Kim, Y. J. Kim, M. E. Kleber, M. J. Knol, A. Kurbasic, M. Lauzon, P. Le, R. Lea, J.-Y. Lee, H. L. Leonard, S. A. Li, X. Li, X. Li, J. Liang, H. Lin, S.-Y. Lin, J. Liu, X. Liu, K. S. Lo, J. Long, L. Lores-Motta, J. Luan, V. Lyssenko, L.-P. Lyytikäinen, A. Mahajan, V. Mamakou, M. Mangino, A. Manichaikul, J. Marten, M. Mattheisen, L. Mavarani, A. F. McDaid, K. Meidtner, T. L. Melendez, J. M. Mercader, Y. Milaneschi, J. E. Miller, I. Y. Millwood, P. P. Mishra, R. E. Mitchell, L. T. Møllehave, A. Morgan, S. Mucha, M. Munz, M. Nakatochi, C. P. Nelson, M. Nethander, C. W. Nho, A. A. Nielsen, I. M. Nolte, S. S. Nongmaithem, R. Noordam, I. Ntalla, T. Nutile, A. Pandit, P. Christofidou, K. Pärna, M. Pauper, E. R. B. Petersen, L. V. Petersen, N. Pitkänen, O. Polašek, A. Poveda, M. H. Preuss, S. Pyarajan, L. M. Raffield, H. Rakugi, J. Ramirez, A. Rasheed, D. Raven, N. W. Rayner, C. Riveros, R. Rohde, D. Ruggiero, S. E. Ruotsalainen, K. A. Ryan, M. Sabater-Lleal, R. Saxena, M. Scholz, A. Sendamarai, B. Shen, J. Shi, J. H. Shin, C. Sidore, C. M. Sitlani, R. C. Slieker, R. A. J. Smit, A. V. Smith, J. A. Smith, L. J. Smyth, L. Southam, V. Steinthorsdottir, L. Sun, F. Takeuchi, D. S. P. Tallapragada, K. D. Taylor, B. O. Tayo, C. Tcheandjieu, N. Terzikhan, P. Tesolin, A. Teumer, E. Theusch, D. J. Thompson, G. Thorleifsson, P. R. H. J. Timmers, S. Trompet, C. Turman, S. Vaccargiu, S. W. van der Laan, P. J. van der Most, J. B. van Klinken, J. van Setten, S. S. Verma, N. Verweij, Y. Veturi, C. A. Wang, C. Wang, L. Wang, Z. Wang, H. R. Warren, W. Bin Wei, A. R. Wickremasinghe, M. Wielscher, K. L. Wiggins, B. S. Winsvold, A. Wong, Y. Wu, M. Wuttke, R. Xia, T. Xie, K. Yamamoto, J. Yang, J. Yao, H. Young, N. A. Yousri, L. Yu, L. Zeng, W. Zhang, X. Zhang, J.-H. Zhao, W. Zhao, W. Zhou, M. E. Zimmermann, M. Zoledziewska, L. S. Adair, H. H. H. Adams, C. A. Aguilar-Salinas, F. Al-Mulla, D. K. Arnett, F. W. Asselbergs, B. O. Åsvold, J. Attia, B. Banas, S. Bandinelli, D. A. Bennett, T. Bergler, D. Bharadwaj, G. Biino, H. Bisgaard, E. Boerwinkle, C. A. Böger, K. Bønnelykke, D. I. Boomsma, A. D. Børglum, J. B. Borja, C. Bouchard, D. W. Bowden, I. Brandslund, B. Brumpton, J. E. Buring, M. J. Caulfield, J. C. Chambers, G. R. Chandak, S. J. Chanock, N. Chaturvedi, Y.-D. I. Chen, Z. Chen, C.-Y. Cheng, I. E. Christophersen, M. Ciullo, J. W. Cole, F. S. Collins, R. S. Cooper, M. Cruz, F. Cucca, L. A. Cupples, M. J. Cutler, S. M. Damrauer, T. M. Dantoft, G. J. de Borst, L. C. P. G. M. de Groot, P. L. De Jager, D. P. V. de Kleijn, H. Janaka de Silva, G. V. Dedoussis, A. I. den Hollander, S. Du, D. F. Easton, P. J. M. Elders, A. H. Eliassen, P. T. Ellinor, S. Elmståhl, J. Erdmann, M. K. Evans, D. Fatkin, B. Feenstra, M. F. Feitosa, L. Ferrucci, I. Ford, M. Fornage, A. Franke, P. W. Franks, B. I. Freedman, P. Gasparini, C. Gieger, G. Girotto, M. E. Goddard, Y. M. Golightly, C. Gonzalez-Villalpando, P. Gordon-Larsen, H. Grallert, S. F. A. Grant, N. Grarup, L. Griffiths, V. Gudnason, C. Haiman, H. Hakonarson, T. Hansen, C. A. Hartman, A. T. Hattersley, C. Hayward, S. R. Heckbert, C.-K. Heng, C. Hengstenberg, A. W. Hewitt, H. Hishigaki, C. B. Hoyng, P. L. Huang, W. Huang, S. C. Hunt, K. Hveem, E. Hyppönen, W. G. Iacono, S. Ichihara, M. A. Ikram, C. R. Isasi, R. D. Jackson, M.-R. Jarvelin, Z.-B. Jin, K.-H. Jöckel, P. K. Joshi, P. Jousilahti, J. W. Jukema, M. Kähönen, Y. Kamatani, K. D. Kang, J. Kaprio, S. L. R. Kardia, F. Karpe, N. Kato, F. Kee, T. Kessler, A. V. Khera, C. C. Khor, L. A. L. M. Kiemeney, B.-J. Kim, E. K. Kim, H.-L. Kim, P. Kirchhof, M. Kivimaki, W.-P. Koh, H. A. Koistinen, G. D. Kolovou, J. S. Kooner, C. Kooperberg, A. Köttgen, P. Kovacs, A. Kraaijeveld, P. Kraft, R. M. Krauss, M. Kumari, Z. Kutalik, M. Laakso, L. A. Lange, C. Langenberg, L. J. Launer, L. Le Marchand, H. Lee, N. R. Lee, T. Lehtimäki, H. Li, L. Li, W. Lieb, X. Lin, L. Lind, A. Linneberg, C.-T. Liu, J. Liu, M. Loeffler, B. London, S. A. Lubitz, S. J. Lye, D. A. Mackey, R. Mägi, P. K. E. Magnusson, G. M. Marcus, P. M. Vidal, N. G. Martin, W. März, F. Matsuda, R. W. McGarrah, M. McGue, A. J. McKnight, S. E. Medland, D. Mellström, A. Metspalu, B. D. Mitchell, P. Mitchell, D. O. Mook-Kanamori, A. D. Morris, L. A. Mucci, P. B. Munroe, M. A. Nalls, S. Nazarian, A. E. Nelson, M. J. Neville, C. Newton-Cheh, C. S. Nielsen, M. M. Nöthen, C. Ohlsson, A. J. Oldehinkel, L. Orozco, K. Pahkala, P. Pajukanta, C. N. A. Palmer, E. J. Parra, C. Pattaro, O. Pedersen, C. E. Pennell, B. W. J. H. Penninx, L. Perusse, A. Peters, P. A. Peyser, D. J. Porteous, D. Posthuma, C. Power, P. P. Pramstaller, M. A. Province, Q. Qi, J. Qu, D. J. Rader, O. T. Raitakari, S. Ralhan, L. S. Rallidis, D. C. Rao, S. Redline, D. F. Reilly, A. P. Reiner, S. Y. Rhee, P. M. Ridker, M. Rienstra, S. Ripatti, M. D. Ritchie, D. M. Roden, F. R. Rosendaal, J. I. Rotter, I. Rudan, F. Rutters, C. Sabanayagam, D. Saleheen, V. Salomaa, N. J. Samani, D. K. Sanghera, N. Sattar, B. Schmidt, H. Schmidt, R. Schmidt, M. B. Schulze, H. Schunkert, L. J. Scott, R. J. Scott, P. Sever, E. J. Shiroma, M. B. Shoemaker, X.-O. Shu, E. M. Simonsick, M. Sims, J. R. Singh, A. B. Singleton, M. F. Sinner, J. G. Smith, H. Snieder, T. D. Spector, M. J. Stampfer, K. J. Stark, D. P. Strachan, L. M. ‘t Hart, Y. Tabara, H. Tang, J.-C. Tardif, T. A. Thanaraj, N. J. Timpson, A. Tönjes, A. Tremblay, T. Tuomi, J. Tuomilehto, M.-T. Tusié-Luna, A. G. Uitterlinden, R. M. van Dam, P. van der Harst, N. Van der Velde, C. M. van Duijn, N. M. van Schoor, V. Vitart, U. Völker, P. Vollenweider, H. Völzke, N. H. Wacher-Rodarte, M. Walker, Y. X. Wang, N. J. Wareham, R. M. Watanabe, H. Watkins, D. R. Weir, T. M. Werge, E. Widen, L. R. Wilkens, G. Willemsen, W. C. Willett, J. F. Wilson, T.-Y. Wong, J.-T. Woo, A. F. Wright, J.-Y. Wu, H. Xu, C. S. Yajnik, M. Yokota, J.-M. Yuan, E. Zeggini, B. S. Zemel, W. Zheng, X. Zhu, J. M. Zmuda, A. B. Zonderman, J.-A. Zwart, 23andMe Research Team, G. C. Partida, VA Million Veteran Program, Y. Sun, DiscovEHR (DiscovEHR and MyCode Community Health Initiative), eMERGE (Electronic Medical Records and Genomics Network), D. Croteau-Chonka, Lifelines Cohort Study, J. M. Vonk, The PRACTICAL Consortium, S. Chanock, L. Le Marchand, Understanding Society Scientific Group, D. I. Chasman, Y. S. Cho, I. M. Heid, M. I. McCarthy, M. C. Y. Ng, C. J. O’Donnell, F. Rivadeneira, U. Thorsteinsdottir, Y. V. Sun, E. S. Tai, M. Boehnke, P. Deloukas, A. E. Justice, C. M. Lindgren, R. J. F. Loos, K. L. Mohlke, K. E. North, K. Stefansson, R. G. Walters, T. W. Winkler, K. L. Young, P.-R. Loh, J. Yang, T. Esko, T. L. Assimes, A. Auton, G. R. Abecasis, C. J. Willer, A. E. Locke, S. I. Berndt, G. Lettre, T. M. Frayling, Y. Okada, A. R. Wood, P. M. Visscher, J. N. Hirschhorn, A saturated map of common genetic variants associated with human height. Nature. 610, 704–712 (2022).

42. B. C. Brown, C. J. Ye, A. L. Price, N. Zaitlen, Transethnic Genetic-Correlation Estimates from Summary Statistics. Am. J. Hum. Genet. 99, 76–88 (2016).

43. R. Wrigley, A. J. Phipps-Green, R. K. Topless, T. J. Major, M. Cadzow, P. Riches, A.-K. Tausche, M. Janssen, L. A. B. Joosten, T. L. Jansen, A. So, J. Harré Hindmarsh, L. K. Stamp, N. Dalbeth, T. R. Merriman, Pleiotropic effect of the ABCG2 gene in gout: involvement in serum urate levels and progression from hyperuricemia to gout. Arthritis Res. Ther. 22, 45 (2020).

44. M. Pilon, G. Leclair, E. Oussaïd, I. St_Jean, M. Jutras, M. Gaulin, I. Mongrain, D. Busseuil, J. L. Rouleau, J. Tardif, M. Dubé, S. de Denus, An association study of *ABCG2* rs2231142 on the concentrations of allopurinol and its metabolites. Clin. Transl. Sci. 15, 2024–2034 (2022).

45. K.-H. Yu, P.-Y. Chang, S.-C. Chang, Y.-H. Wu-Chou, L.-A. Wu, D.-P. Chen, F.-S. Lo, J.-J. Lu, A comprehensive analysis of the association of common variants of ABCG2 with gout. Sci. Rep. 7, 9988 (2017).

46. L. M. Polfus, B. F. Darst, H. Highland, X. Sheng, M. C. Y. Ng, J. E. Below, L. Petty, S. Bien, X. Sim, W. Wang, P. Fontanillas, Y. Patel, 23andMe Research Team, DIAMANTE Hispanic/Latino Consortium, MEta-analysis of type 2 DIabetes in African Americans Consortium, M. Preuss, C. Schurmann, Z. Du, Y. Lu, S. K. Rhie, J. M. Mercader, T. Tusie-Luna, C. González-Villalpando, L. Orozco, C. N. Spracklen, B. E. Cade, R. A. Jensen, M. Sun, Y. Y. Joo, P. An, L. R. Yanek, L. F. Bielak, S. Tajuddin, A. Nicolas, G. Chen, L. Raffield, X. Guo, W.-M. Chen, G. N. Nadkarni, M. Graff, R. Tao, J. S. Pankow, M. Daviglus, Q. Qi, E. A. Boerwinkle, S. Liu, L. S. Phillips, U. Peters, C. Carlson, L. R. Wikens, L. L. Marchand, K. E. North, S. Buyske, C. Kooperberg, R. J. F. Loos, D. O. Stram, C. A. Haiman, Genetic discovery and risk characterization in type 2 diabetes across diverse populations. HGG Adv. 2, 100029 (2021).

47. J. Chen, M. Sun, A. Adeyemo, F. Pirie, T. Carstensen, C. Pomilla, A. P. Doumatey, G. Chen, E. H. Young, M. Sandhu, A. P. Morris, I. Barroso, M. I. McCarthy, A. Mahajan, E. Wheeler, C. N. Rotimi, A. A. Motala, Genome-wide association study of type 2 diabetes in Africa. Diabetologia. 62, 1204–1211 (2019).

48. F. Rajabli, G. W. Beecham, H. C. Hendrie, O. Baiyewu, A. Ogunniyi, S. Gao, N. A. Kushch, M. Lipkin-Vasquez, K. L. Hamilton-Nelson, J. I. Young, D. M. Dykxhoorn, K. Nuytemans, B. W. Kunkle, L. Wang, F. Jin, X. Liu, B. E. Feliciano-Astacio, Alzheimer’s Disease Sequencing Project, Alzheimer’s Disease Genetic Consortium, G. D. Schellenberg, C. L. Dalgard, A. J. Griswold, G. S. Byrd, C. Reitz, M. L. Cuccaro, J. L. Haines, M. A. Pericak-Vance, J. M. Vance, A locus at 19q13.31 significantly reduces the ApoE ε4 risk for Alzheimer’s Disease in African Ancestry. PLOS Genet. 18, e1009977 (2022).

49. J. D. Martin-Rufino, N. Castano, M. Pang, E. I. Grody, S. Joubran, A. Caulier, L. Wahlster, T. Li, X. Qiu, A. M. Riera-Escandell, G. A. Newby, A. Al’Khafaji, S. Chaudhary, S. Black, C. Weng, G. Munson, D. R. Liu, M. W. Wlodarski, K. Sims, J. H. Oakley, R. M. Fasano, R. J. Xavier, E. S. Lander, D. E. Klein, V. G. Sankaran, Massively parallel base editing to map variant effects in human hematopoiesis. Cell. 186, 2456–2474.e24 (2023).

50. O. Weissbrod, F. Hormozdiari, C. Benner, R. Cui, J. Ulirsch, S. Gazal, A. P. Schoech, B. van de Geijn, Y. Reshef, C. Márquez-Luna, L. O’Connor, M. Pirinen, H. K. Finucane, A. L. Price, Functionally informed fine-mapping and polygenic localization of complex trait heritability. Nat. Genet. 52, 1355–1363 (2020).

51. S. Rao, Y. Yao, D. E. Bauer, Editing GWAS: experimental approaches to dissect and exploit disease-associated genetic variation. Genome Med. 13, 41 (2021).

52. S. B. Gabriel, S. F. Schaffner, H. Nguyen, J. M. Moore, J. Roy, B. Blumenstiel, J. Higgins, M. DeFelice, A. Lochner, M. Faggart, S. N. Liu-Cordero, C. Rotimi, A. Adeyemo, R. Cooper, R. Ward, E. S. Lander, M. J. Daly, D. Altshuler, The Structure of Haplotype Blocks in the Human Genome. Science. 296, 2225–2229 (2002).

53. D. E. Reich, M. Cargill, S. Bolk, J. Ireland, P. C. Sabeti, D. J. Richter, T. Lavery, R. Kouyoumjian, S. F. Farhadian, R. Ward, E. S. Lander, Linkage disequilibrium in the human genome. Nature. 411, 199–204 (2001).

54. M. M. Hsieh, J. E. Everhart, D. D. Byrd-Holt, J. F. Tisdale, G. P. Rodgers, Prevalence of Neutropenia in the U.S. Population: Age, Sex, Smoking Status, and Ethnic Differences. Ann. Intern. Med. 146, 486 (2007).

55. M. A. Nalls, J. G. Wilson, N. J. Patterson, A. Tandon, J. M. Zmuda, S. Huntsman, M. Garcia, D. Hu, R. Li, B. A. Beamer, K. V. Patel, E. L. Akylbekova, J. C. Files, C. L. Hardy, S. G. Buxbaum, H. A. Taylor, D. Reich, T. B. Harris, E. Ziv, Admixture Mapping of White Cell Count: Genetic Locus Responsible for Lower White Blood Cell Count in the Health ABC and Jackson Heart Studies. Am. J. Hum. Genet. 82, 81–87 (2008).

56. N. McCormick, N. Lu, C. Yokose, A. D. Joshi, S. Sheehy, L. Rosenberg, E. T. Warner, N. Dalbeth, T. R. Merriman, K. G. Saag, Y. Zhang, H. K. Choi, Racial and Sex Disparities in Gout Prevalence Among US Adults. JAMA Netw. Open. 5, e2226804 (2022).

57. A. J. Griswold, K. Celis, P. L. Bussies, F. Rajabli, P. L. Whitehead, K. L. Hamilton-Nelson, G. W. Beecham, D. M. Dykxhoorn, K. Nuytemans, L. Wang, O. K. Gardner, D. A. Dorfsman, E. H. Bigio, M. M. Mesulam, S. Weintraub, C. Geula, M. Gearing, E. McGrath-Martinez, C. L. Dalgard, W. K. Scott, J. L. Haines, M. A. Pericak-Vance, J. I. Young, J. M. Vance, Increased APOE ε4 expression is associated with the difference in Alzheimer’s disease risk from diverse ancestral backgrounds. Alzheimers Dement. J. Alzheimers Assoc. 17, 1179–1188 (2021).

58. J. Pulley, E. Clayton, G. R. Bernard, D. M. Roden, D. R. Masys, Principles of human subjects protections applied in an opt-out, de-identified biobank. Clin. Transl. Sci. 3, 42–48 (2010).

59. Sanger Imputation Service, (available at https://imputation.sanger.ac.uk/?about=1#referencepanels).

60. 1000 Genomes Project Consortium, A. Auton, L. D. Brooks, R. M. Durbin, E. P. Garrison, H. M. Kang, J. O. Korbel, J. L. Marchini, S. McCarthy, G. A. McVean, G. R. Abecasis, A global reference for human genetic variation. Nature. 526, 68–74 (2015).

61. N. Patterson, A. L. Price, D. Reich, Population Structure and Eigenanalysis. PLoS Genet. 2, e190 (2006).

62. L. Bastarache, Using Phecodes for Research with the Electronic Health Record: From PheWAS to PheRS. Annu. Rev. Biomed. Data Sci. 4, 1–19 (2021).

63. X.-M. T. Nguyen, S. B. Whitbourne, Y. Li, R. M. Quaden, R. J. Song, H.-N. A. Nguyen, K. Harrington, L. Djousse, J. V. V. Brewer, J. Deen, S. Muralidhar, R. B. Ramoni, K. Cho, J. P. Casas, P. S. Tsao, J. M. Gaziano, the VA Million Veteran Program, Data Resource Profile: Self-reported data in the Million Veteran Program: survey development and insights from the first 850_736 participants. Int. J. Epidemiol. 52, e1–e17 (2023).

64. T. W. Winkler, F. R. Day, D. C. Croteau-Chonka, A. R. Wood, A. E. Locke, R. Mägi, T. Ferreira, T. Fall, M. Graff, A. E. Justice, J. Luan, S. Gustafsson, J. C. Randall, S. Vedantam, T. Workalemahu, T. O. Kilpeläinen, A. Scherag, T. Esko, Z. Kutalik, I. M. Heid, R. J. F. Loos, Genetic Investigation of Anthropometric Traits (GIANT) Consortium, Quality control and conduct of genome-wide association meta-analyses. Nat. Protoc. 9, 1192–1212 (2014).

65. C. C. Chang, C. C. Chow, L. C. Tellier, S. Vattikuti, S. M. Purcell, J. J. Lee, Second-generation PLINK: rising to the challenge of larger and richer datasets. GigaScience. 4 (2015), doi:10.1186/s13742-015-0047-8.

66. C. Wallace, C. Giambartolomei, V. Plagnol, coloc: Colocalisation Tests of Two Genetic Traits (2023), (available at https://cran.r-project.org/web/packages/coloc/index.html).

67. C. J. Willer, Y. Li, G. R. Abecasis, METAL: fast and efficient meta-analysis of genomewide association scans. Bioinforma. Oxf. Engl. 26, 2190–2191 (2010).

68. W. McLaren, L. Gil, S. E. Hunt, H. S. Riat, G. R. S. Ritchie, A. Thormann, P. Flicek, F. Cunningham, The Ensembl Variant Effect Predictor. Genome Biol. 17 (2016), doi:10.1186/s13059-016-0974-4.

69. X. Wang, I.-E. Nogues, M. Liu, T. Chen, X. Xiong, C.-L. Bonzel, H. Zhang, C. Hong, K. Dahal, L. Costa, J. M. Gaziano, S. C. Kim, Y.-L. Ho, K. Cho, T. Cai, K. P. Liao, Differential Associations of Interleukin 6 Receptor Variant Across Genetic Ancestries and Implications for Targeted Therapies (2022), p. 2022.09.24.22280325, doi:10.1101/2022.09.24.22280325.

