## Supplementary Note for "Diversity and Scale: Genetic Architecture of 2,068 Traits in the VA Million Veteran Program"

**Supplementary Information for the paper**

#### Introduction

This supplementary document provides additional information and details about the study that cannot be included in the main paper due to space constraints. Here we include details about the study design, the statistical methods used, and the results of the analysis, as well as any additional tables or figures that provide further insight into the study findings.

#### Phenotypes

The initial phenotype data were accessed through the Centralized Interactive Phenomics Resource (CIPHER, https://phenomics.va.ornl.gov/), which consists of electronic health record (EHR)-based phenotypes derived from various sources, including EHR diagnosis codes, clinical laboratory tests, and survey questionnaire responses. In this study, we utilized phenotypes from all three sources to examine the associations with genetic variants.

**Phenotypes from ICD codes**

The binary phenotypes from the electronic health records (EHRs) were defined using phecodes as previously described in Wei et al. (<https://doi.org/10.1371/journal.pone.0175508>). Each phecode represents a group of International Classification of Diseases (ICD) codes clinically relevant for a particular phenotype. Using this approach, all ICD codes for all Veterans in the MVP cohort were extracted and assigned a phecode-defined phenotype. ICD-9 and ICD-10 codes were mapped to 1,876 phecodes. For each phecode, participants with at least two phecode-mapped ICD-9 or ICD-10 codes were defined as cases, while those with no instances of a phecode-mapped ICD-9 or ICD-10 code were defined as controls. Based on our previous simulation studies of EHR data, populations where the phecode comprises fewer than 500 cases are more likely to produce spurious results, so we applied this threshold in four ancestry groups: African ancestry (AFR), American ancestry (AMR), European ancestry (EUR), and Asian ancestry (ASN).

**Phenotype from Laboratory measurements**

We defined quantitative phenotypes based on the laboratory measurements collected during outpatient and inpatient visits. For quantitative traits, we calculated the mean, median, and max values across all visits for each participant and analyzed only the resulting phenotype. Only quantitative traits with data for more than 1,000 individuals were included in the analyses. The remaining 69 laboratory measurements that passed quality control were normalized using a rank-based inverse-normal transformation. To remove extreme outliers, we remove values greater than six standard deviations from the trait mean.

**Survey questionnaire**

The two surveys (questionnaires) for MVP, as noted previously, were designed to augment data contained in each participant's electronic health record. As with other study activities and all study materials sent to participants, these documents were approved by the VA Central IRB. As participants are enrolled, informed consent and HIPAA authorization forms are scanned by field site staff and sent to the CERC to be checked for accuracy and completeness, and the data are entered in GenISIS. Conceptually, the MVP Baseline Survey was designed to collect information regarding demographics, family pedigree, health status, lifestyle habits, military experience, medical history, family history of specific illnesses, and physical features. The MVP Lifestyle Survey contains questions from validated instruments in domains selected to provide information on sleep and exercise habits, environmental exposures, dietary habits, and sense of well-being. We selected surveys with either binary responses (yes/no) or quantitative responses (such as height, weight, age at smoking onset, and age at menopause). To align with laboratory measurements, we normalized all quantitative traits using a rank-based inverse-normal transformation.

#### Ancestry assignment

To estimate ancestry, we obtained a reference dataset from the 1000 Genomes Project and used the smartpca module in the EIGENSOFT (*1*) package to project the PC loadings from a group of unrelated individuals in the reference dataset. We merged this dataset with the MVP dataset and ran smartpca to project the PCA loadings from the reference dataset. We trained a random forest classifier using continental ancestry meta-data based on the top 10 principal components from the reference training data to define genetically inferred ancestry. We then applied this random forest to the predicted MVP PCA data and assigned ancestries to individuals with a probability greater than 50%. Those with a probability less than 50% for any particular ancestry group were excluded from the study. Figure X shows the PCA projection of MVP participants on the 1000 Genomes reference panel. Lastly, we selected PC1-10 to adjust for our and scree plot in Figure shows the variance explained by each PC.

Additionally, we also utilized harmonized ancestry and race/ethnicity (HARE)(59) population groups and implemented the same quality control (QC) criteria. The HARE group's GWAS summary statistics will also be readily available via the dbGAP portal.

#### Optimization of SAIGE

This study aimed to conduct a series of genome-wide association scans (GWAS) using a curated set of 2,068 traits. The GWAS was conducted using the linear mixed model method implemented in the SAIGE R package, which was optimized to run on the OLCF Summit infrastructure. We optimized the use of the SAIGE package by running it on the OLCF Summit infrastructure, which used the power of GPUs to speed up the matrix computation involved in building the genetic relationship matrix. This optimization allowed us to distribute the matrix operations evenly among a number of GPUs depending on the size of the genetic relationship matrix. For example, for the European cohort, we were able to use eight high-memory GPUs, while for the Asian cohort, we only needed 1 GPU. The use of GPUs significantly improved the execution time of the analysis, allowing us to quickly and efficiently conduct the GWAS on a large number of phenotypes (fig S).

#### Creation of reference panels

LD information is crucial in the post-GWAS analysis of summary statistics (*2*). MVP participants were genotyped on a custom chip, including a small number of population-specific variants not present in the external reference panel, such as the 1000 Genomes project (*2*, *3*). Estimating LD among SNPs in a genomic region is critical for GWAS analysis, and using a reference panel with a population used for GWAS can improve the accuracy of LD estimation. External reference panels may not be representative of the population being studied, which can lead to biased results. Hence, we created a reference panel representing the population group using the individuals level genetic data from MVP participants from each population group. In our study, we create three different reference panels to use for different analyses:

A) **Identification of independent loci and lead variants**
   In order to ensure the accuracy of the LD structure, we selected 5000 individuals at random from each of the groups included in the study: African American, Admixed American, East Asian, and European. We then used the PLINK 1.9 software to filter the data, keeping only SNPs with a MAC>20 in each ancestral group. Afterward, we combined all the individual files to create a multi-population reference panel of 20,000 MVP participants. This panel served as a reference for clumping and thresholding to identify independent loci and lead variants from a meta-analysis of summary statistics across multiple populations. By customizing our approach to the study population, we could effectively use the LD structure for GWAS.
B) **LDSC LD score**
   We used LDSC (Linkage Disequilibrium Score Regression) to calculate each trait's SNP heritability and genetic correlation based on the GWAS summary statistics. To generate ancestry-specific LD scores, we followed the steps outlined in this resource (https://github.com/bulik/ldsc/wiki/LD-Score-Estimation-Tutorial). First, we utilized the reference panel developed for each ancestry group in (a), then we subsetted the data to the HapMAP3 SNP list provided by the LDSC developer team. Finally, we used the genotype data to compute the ancestry-specific LD scores with the LDSC software. We repeated these steps for each ancestry group to obtain the respective ancestry-specific LD scores.
C) **Fine-mapping**
   To eliminate artifacts resulting from an LD mismatch between the meta-analyzed GWAS results and any reference panel, we fine-mapped the population-specific GWAS results separately for each significant locus-phenotype combination using matched reference matrices for each of the population groups AFR, AMR, EAS, and EUR ancestries. We prepared the matched matrices for each combination by filtering 1) the genotype data for variants at the locus and 2) individuals included in the population-specific GWAS for the phenotype. We excluded indels and multiallelic sites to avoid potential errors in alignment or strand flipping. After filtering, we calculated all pairwise correlations between the remaining variants using the “--r square --keep-allele-order” options in PLINK v1.9(*4*). Because PLINK uses hard-called genotypes to calculate LD between variants, but our imputation process generated dosage-based genotypes, it was impossible to calculate correlations between specific pairs of variants using PLINK’s default thresholds for hard-call conversion. This occurred both when there were no individuals with convertible genotypes for a given variant and when the sets of individuals with convertible genotypes for distinct variants did not overlap. Because the fine-mapping process cannot tolerate missing correlations in the LD matrix, for any matrix with missing correlations, we iteratively removed the variant with the most missing values until the matrix was complete.

#### Comparison of our fine-mapping results with other biobank-based analyses

Compared to other large-scale fine-mapping experiments(*5*–*7*), our analysis is the biggest in terms of the number of phenotypes mapped and signals identified, and only the analysis by Kanai et al.^1^ is more extensive in terms of samples included.  We borrowed much from Kanai et al. when designing our approach to fine-mapping, though we chose to deviate from them in several respects, primarily in how we defined loci, in the maximum number of signals we attempted to map at each locus, and in what fine-mapping methods we used.

Regarding locus definition, we wanted to minimize the size of each locus while ensuring that we did not arbitrarily crop the pattern of association and create false positive signals. Though our custom approach resulted in some very large loci, including several greater than 10 MB at the previously known Duffy/*DARC* locus on chromosome 1(*8*), in general, it was successful; it allowed us to fine-map 99% of the loci in all four ancestries and 99.7% in at least one ancestry. The key shortcoming of our locus-defining scheme is that it can create overlapping loci, which can and did result in double-counted signals; 115 signals (0.2% of signals) were double-counted. As far as we know, Kanai et al. did not provide fine-mapping completion statistics for their approach, however, their locus-defining scheme should have excluded the possibility of double-counted signals.

At a summary level, our fine-mapping results were broadly similar to those obtained by Kanai et al., except that we obtained more-precise credible sets. Across the three biobanks they analyzed, between 34 and 37% of the signals Kanai et al. mapped contained five or fewer variants. at the same time, 54% of our merged credible sets were fine-mapped to the same precision. We observed an enrichment of rarer variants among more precisely mapped credible sets (**fig. S11**) that could explain this precision difference. Though Kanai et al. applied inconsistent frequency-based variant filters across their cohorts, in general, it appears that we retained more rare variants in our analysis. Since we observed important coding (**Fig. 3D**) and noncoding (**fig. S11**) functional enrichments among our precisely mapped credible sets, the inclusion of these rarer variants was robust and an advantage of our approach. Apart from allele frequencies, differences in which phenotypes were fine-mapped could also explain the precision gap, though this possibility is less likely. We examined the precision of our fine-mapped signals across the five phenotype categories we examined, and even in the lowest category we fine-mapped 41% of the signals to five or fewer variants (**fig. S5**). Sub category-level differences in precision rates could still account for the precision difference, as could differences in LD matrix construction, population composition, and fine-mapping methods. However, more work would be needed to investigate these possibilities.

#### Heterogeneity Analysis

We developed a statistical testing framework for assessing the heterogeneity of genetic associations with multiple phenotypes across different ancestry groups. The goal is to simultaneously test whether the effects of genetic variants on the phenotypes are homogeneous across all groups or if heterogeneity is present in at least one group. The framework uses various statistical models, including linear and logistic regression, to model the relationships between genetic variants, adjustment covariates, and phenotypes.

**1. Problem Setup**

Let $J$ be the number of ancestry groups considered for testing heterogeneity. For each subject $i$ from each group $j$, we observe a $K$-dimensional vector of outcomes $Y_{i,j}=\left( Y_{i,j,1},Y_{i,j,2},\ldots,Y_{i,j,K} \right)^{T}$, a vector of adjustment covariates $X_{i,j}$ with demographic variables, and an $L$-dimensional vector of SNPs $A_{i,j}=$ $\left( A_{i,j,1},A_{i,j,2},\ldots,A_{i,j,L} \right)^{T}$. Within each ancestry group $j$, the outcome $Y_{i,j,k}$ is modeled against each SNP $A_{i,j,\mathcal{l}}$ and adjustment covariates $X_{i,j}$. For continuous outcomes, we consider the linear model:

$$Y_{i,j,k}=\beta_{j,\mathcal{l},k}A_{i,j,\mathcal{l}}+\gamma_{j,\mathcal{l},k}^{T}X_{i,j}+\epsilon_{i,j,k}, \text{ where } \epsilon_{i,j,k}\sim N\left( 0,\sigma_{j,k}^{2} \right)$$

while for binary outcomes, we use the logistic model:

$$P\left( Y_{i,j,k}=1\mid A_{i,j,\mathcal{l}},X_{i,j} \right)=expit\left( \beta_{j,\mathcal{l},k}A_{i,j,\mathcal{l}}+\gamma_{j,\mathcal{l},k}^{T}X_{i,j} \right),$$

where $expit(a)=e^{a}/\left( 1+e^{a} \right)$. For each pair of phenotype $k$ and SNP $\mathcal{l}$, let $\beta_{j,\mathcal{l},k}=\mu_{\mathcal{l},k}+\alpha_{j,\mathcal{l},k}$ where $\mu_{\mathcal{l},k}$ denotes the mean association between outcome $k$ and SNP $\mathcal{l}$ across all ancestry groups and $\alpha_{j,\mathcal{l},k}$ reflects the heterogeneity in effect from group $j$. Define the true non-null sets $\mathcal{S}_{\mu}=\left\{ (\mathcal{l},k):\mu_{\mathcal{l}k}\neq0 \right\}$ and $\mathcal{S}_{\alpha}=\left\{ (\mathcal{l},k):\alpha_{j,\mathcal{l}k}\neq0 \right.$ for some $\left. j \right\}$. We assume that approximately: $\mathcal{S}_{\alpha}\subseteq\mathcal{S}_{\mu}$ or $\mathcal{S}_{\alpha}$ and $\mathcal{S}_{\mu}$ are very similar. For $(\mathcal{l},k)\in\{1,\ldots,L\}\times\{1,\ldots,K\}$, we are interested in simultaneously testing:

$$\alpha_{1,\mathcal{l}k}=\cdots=\alpha_{J,\mathcal{l}k}=0\text{ v.s. }\alpha_{j,\mathcal{l}k}\neq0\text{ for some }j\in\{1,\ldots,J\}.$$

with the false discovery rate (FDR) controlled below some level $\eta$ (e.g., $\eta=0.1$ ):

$$E\left[ \frac{\#\text{ of false discovery }}{\text{ Total }\#\text{ of discovery }} \right]\leq\eta\text{. }$$

**2. Test Statistics**

We first construct the effect estimator $\hat{\beta}_{j,\mathcal{l},k}$ for each phenotype $k$ against each SNP $\mathcal{l}$ on each group $j$ and estimate its asymptotic variance $\hat{\sigma}_{j,\mathcal{l},k}$ using the standard score test approach. Then we introduce the mean effect auxiliary statistic constructed as the inverse-variance weighted average across ancestry groups:

$$\hat{\mu}_{\mathcal{l},k}=\frac{\sum_{j=1}^{J} \left( \hat{\sigma}_{j,\mathcal{l},k} \right)^{-2}\hat{\beta}_{j,\mathcal{l},k}}{\sum_{j=1}^{J} \left( \hat{\sigma}_{j,\mathcal{l},k} \right)^{-2}},\text{ which follows }N\left( 0,\frac{1}{\sum_{j=1}^{J} \left( \hat{\sigma}_{j,\mathcal{l},k} \right)^{-2}} \right)\text{ when }\beta_{1,\mathcal{l},k}=\cdots=\beta_{J,\mathcal{l},k}=0,$$

and the heterogeneity test statistic as a quadratic form of the estimated heterogeneity effects:

$$\hat{T}_{\mathcal{l},k}=\sum_{j=1}^{J} \left( \hat{\beta}_{j,\mathcal{l},k}-J^{-1}\sum_{j^{'}=1}^{J} \hat{\beta}_{j^{'},\mathcal{l},k} \right)^{2},\text{ which follows }\sum_{j=1}^{J} \lambda_{j}\chi_{1(j)}^{2}\text{ when }\beta_{1,\mathcal{l},k}=\cdots=\beta_{J,\mathcal{l},k}.$$

Here $\chi_{1(j)}^{2},\ldots,\chi_{1(J)}^{2}$ are $J$ mutually independent chi-square random variables with degree of freedom 1 , and $\lambda_{j}$ 's are estimated by calculating the eigenvalues of the $J\times J$ empirical covariance matrix:

$$\hat{\Sigma}_{\mathcal{l},k}=\frac{\sum_{j} \left( \hat{\sigma}_{j,\mathcal{l},k} \right)^{2}}{J^{2}}\mathbf{1}-\left[ diag\left( \frac{\left( \hat{\sigma}_{j,\mathcal{l},k} \right)^{2}}{J} \right) \right]_{j=1}^{J}\mathbf{1}_{J}-\mathbf{1}_{J}\left[ diag\left( \frac{\left( \hat{\sigma}_{j,\mathcal{l},k} \right)^{2}}{J} \right) \right]_{j=1}^{J}+\left[ diag\left( \left( \hat{\sigma}_{j,\mathcal{l},k} \right)^{2} \right) \right]_{j=1}^{J},$$

where $\mathbf{1}_{J}$ is a $J\times J$ matrix of all ones. The $p$-value of $\hat{T}_{\mathcal{l},k}$, denoted as $\hat{p}_{\alpha,\mathcal{l},k}$, characterizes significance of the across-group heterogeneity of the association between phenotype $k$ and SNP $\mathcal{l}$, and is calculated using CompQuadForm package in $\mathbf{R}(5)\mathbf{.}$ Meanwhile, we extract the $p$-value of the mean effect statistics $\hat{\mu}_{\mathcal{l},k}$, denoted as $\hat{p}_{\mu,k,\mathcal{l}}$, as a guiding auxiliary information to enhance the testing power. It is not hard to show that $\hat{\mu}_{\mathcal{l},k}$ is asymptotically independent with $\hat{\beta}_{j,\mathcal{l},k}-J^{-1}\sum_{j^{'}=1}^{J} \hat{\beta}_{j^{'},\mathcal{l},k}$ for every $j=1,2,\ldots,J$, and, thus, is asymptotically independent with $\hat{T}_{\mathcal{l},k}$. This grants the validity of using each $\hat{p}_{\mu,k,\mathcal{l}}$ as a side information to re-weight $\hat{p}_{\alpha,k,\mathcal{l}}$, which is an important step to be introduced in the next section.

**3. Weights Construction**

We aim at taking $\hat{p}_{\mu,\mathcal{l},k}$ as an auxiliary information to assign weights to $\hat{p}_{\alpha,\mathcal{l},k}$, based upon the prior assumption that the non-null set of heterogeneity $\mathcal{S}_{\alpha}$ is close to that of the mean effects $\mathcal{S}_{\mu}$. Asymptotic independence between $\hat{p}_{\mu,\mathcal{l},k}$ and $\hat{p}_{\alpha,\mathcal{l},k}$ warrants the validity of this strategy and the large effective sample size for the overall effect $\hat{p}_{\mu,\mathcal{l},k}$ makes it a precise enough information to enhance the power of the heterogeneity effects testing. Inspired by recent literature of adaptive multiple testing(*9*, *10*) that leverages auxiliary information to improve the power over the standard Benjamini Hochberg (BH) procedure (*11*), we propose the following steps to derive $\hat{p}_{\mu,\mathcal{l},k}$ into proper weights of the testing $p$-values $\hat{p}_{\alpha,\mathcal{l},k}$ :

1 Calculate $Z_{\alpha,\mathcal{l},k}=I\left( \hat{p}_{\alpha,\mathcal{l},k}<\tau_{\alpha} \right)$ where $\tau_{\alpha}$ is some pre-specified cutoff parameter. Practically, one can either fix $\tau_{\alpha}$ as some small value like ${10}^{-5}$ or specify it empirically, e.g. choosing $\tau_{\alpha}$ as the $p$-value cutoff returned from the BH procedure on $\left\{ \hat{p}_{\alpha,k}:k=1,2,\ldots,K \right\}$ with level $0.5$ (Cai et al., 2020).
2 Implement a logistic regression on

$$Z_{\alpha,\mathcal{l},k}\sim a\cdot m \left\{ logit\left( 1-\hat{p}_{\mu,\mathcal{l},k} \right),12 \right\}+b_{k},$$

where $logit(x)=log\{x/(1-x)\},a$ is a regression coefficient and $b_{k}$ represents coefficient for the phenotype $k$. Since $\hat{p}_{\mu,\mathcal{l},k}$ could be extremely close to zero, we use logit-transformation and threshold on $\hat{p}_{\mu,\mathcal{l},k}$ to make the weighting model more stable. Let the fitted coefficients be $\hat{a}$ and $\hat{b}_{k}$ for $k=$ $1,2,\ldots,K$. We set the weight for $\hat{p}_{\alpha,\mathcal{l},k}$ as

$$\hat{\pi}_{\mathcal{l},k}=m \left\{ 1-\frac{1-exp\left\{ \hat{a}\cdot m \left\{ logit\left( 1-\hat{p}_{\mu,\mathcal{l},k} \right),12 \right\}+\hat{b}_{k} \right\}}{1-\tau_{\alpha}},0 \right\}.$$

3 Standardize $\hat{\pi}_{\mathcal{l},k}$ and obtain the final weights $\hat{q}_{\mathcal{l},k}$ through: $\hat{q}_{\mathcal{l},k}=KL\left( \sum_{k=1}^{K} \sum_{\mathcal{l}=1}^{L} \hat{\pi}_{\mathcal{l},k} \right)^{-1}\hat{\pi}_{\mathcal{l},k}$.

Construction of $\hat{\pi}_{\mathcal{l},k}$ is inspired by the optimal bayesian decision rule used in Cai et al.(*10*). Since we do not observe the true set with heterogeneity effects $\mathcal{S}_{\alpha}$, we use $Z_{\alpha,\mathcal{l},k}=I\left( \hat{p}_{\alpha,\mathcal{l},k}<\tau_{\alpha} \right)$ as a surrogate for the presence of heterogeneity on phenotype $k$ and SNP $\mathcal{l}$, and perform regression on it against the auxiliary information to derive the weight $\hat{\pi}_{\mathcal{l},k}$. Since $Z_{\alpha,\mathcal{l},k}$ depends on the testing $p$-value $\hat{p}_{\alpha,\mathcal{l},k}$, the final FDR control procedure described below is modified correspondingly to ensure validity. Our third step of standardizing $\hat{\pi}_{\mathcal{l},k}$ is also used to protect the validity and FDR control.

**4. Adaptive FDR Control**

Finally, we weight and adjust the heterogeneity testing $p$-values as $\hat{p}_{\alpha,\mathcal{l},k}^{q}=min\left\{ \hat{p}_{\alpha,\mathcal{l},k}/\hat{q}_{\mathcal{l},k},1 \right\}$ for each pair of $\mathcal{l}$ and $k$, and implement the following algorithm for discovery with FDR control of level $\eta$.

**5. Computational Dimensionality Reduction**

Considering the ultra-high-dimensionality of the genetic variants and phenotypes, we use a simple strategy that can reduce the burden of computation and storage while maintaining little loss on power. At the Algorithm 1 Adaptive multiple testing with FDR level $\eta$.

1: Find $\hat{r}=max\left\{ r\geq1:\hat{p}_{\alpha,\mathcal{l},k}^{q}\leq\{r\eta/(KL)\}\wedge\tau_{\alpha} \right.$ for at least $r$ many $\left. \hat{p}_{\alpha,\mathcal{l},k}^{q} \right\}$;

2: Reject null hypothesis with $\hat{p}_{\alpha,\mathcal{l},k}^{q}\leq\{\hat{r}\eta/(KL)\}\wedge\tau_{\alpha}$ for a total of $\hat{r}$ rejections.

beginning, we still use score test to derive the beta coefficient $\hat{\beta}_{j,\mathcal{l},k}$, its asymptotic variance $\hat{\sigma}_{j,\mathcal{l},k}$, and $p$-value $\hat{p}_{\beta,j,\mathcal{l},k}$ for each $j,\mathcal{l}$ and $k$. Then we introduce a modified multiple testing procedure requiring less resource on computation and storage in Algorithm 2.

Algorithm 2 Computational Dimensionality Reduction.

1: For each pair $\mathcal{l}$ and $k$, if $\min\left\{ \hat{p}_{\beta,j,\mathcal{l},k}:j=1,2,\ldots,J \right\}>\tau_{\beta}$, skip the steps of constructing $\hat{\mu}_{\mathcal{l},k},\hat{T}_{\mathcal{l},k}$ and simply set $\hat{p}_{\mu,\mathcal{l},k}=\hat{p}_{\alpha,\mathcal{l},k}=1$; otherwise, computing $\hat{\mu}_{\mathcal{l},k},\hat{T}_{\mathcal{l},k},\hat{p}_{\mu,\mathcal{l},k}$ and $\hat{p}_{\alpha,\mathcal{l},k}$ using the same way described above.

2: Implement the same weight construction procedure with only the pairs satisfying $\hat{p}_{\mu,\mathcal{l},k},\hat{p}_{\alpha,\mathcal{l},k}\neq1$;

3: Implement Algorithm 1 on the union set of re-weighted $p$-values and the $p$-values set as 1 in the first step, for multiple testing with FDR control.

In Algorithm $2,\tau_{\beta}$ is some pre-specified small threshold parameters set as ${10}^{-6}$ in our case. The smaller $\tau_{\beta}$ is, the less computation and storage resource Algorithm 2 will require but there will be a high risk of losing power if $\tau_{\beta}$ is excessively small. Inspired by Liu et al. (2022)(*12*), Algorithm 2 preserves validity because it only makes conservative changes by setting many heterogeneity $p$-values as 1 and still adjusts for the total number of hypotheses as $KL$ in the weighting and FDR control procedures. Meanwhile, the screening procedure, i.e., Step 1 in Algorithm 2 may not impact power since it only excludes pairs of SNP and phenotype with apparently non-significant heterogeneity effects.

#### Tables

All the supplementary tables can be downloaded from <https://tinyurl.com/mww86dn8>.

#### Figures

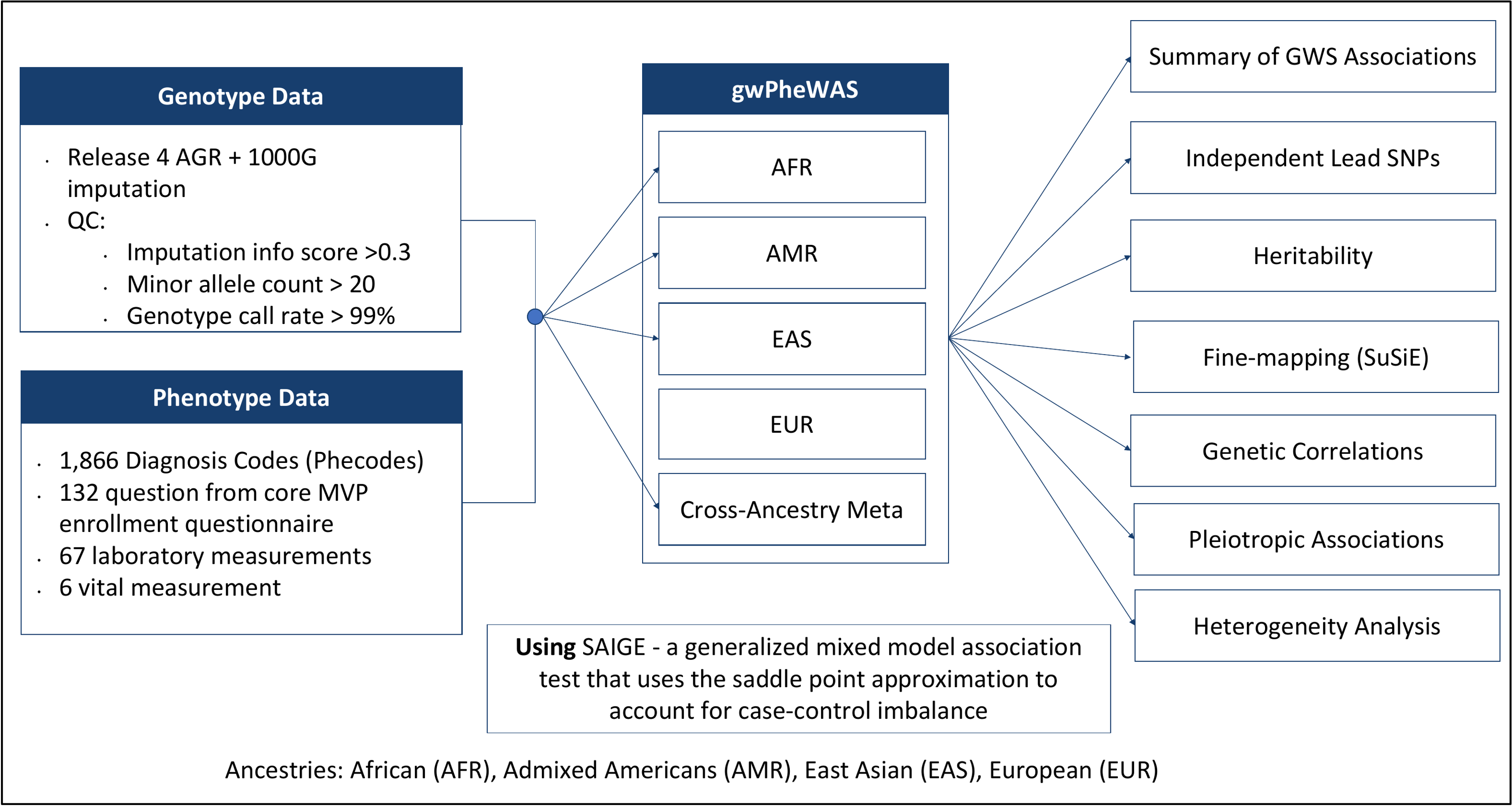

**Figure S1.** Study design overview.

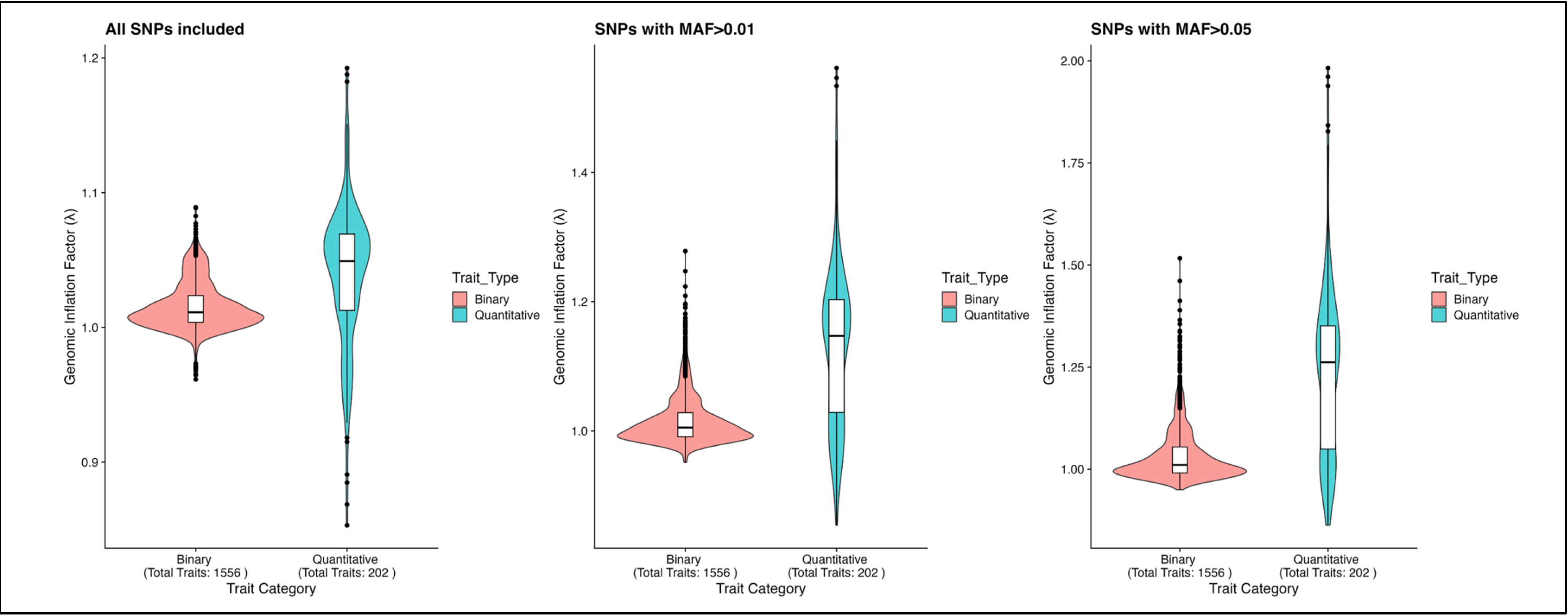

**Figure S2.** Distribution of genomic inflation factor for meta-analyzed traits.

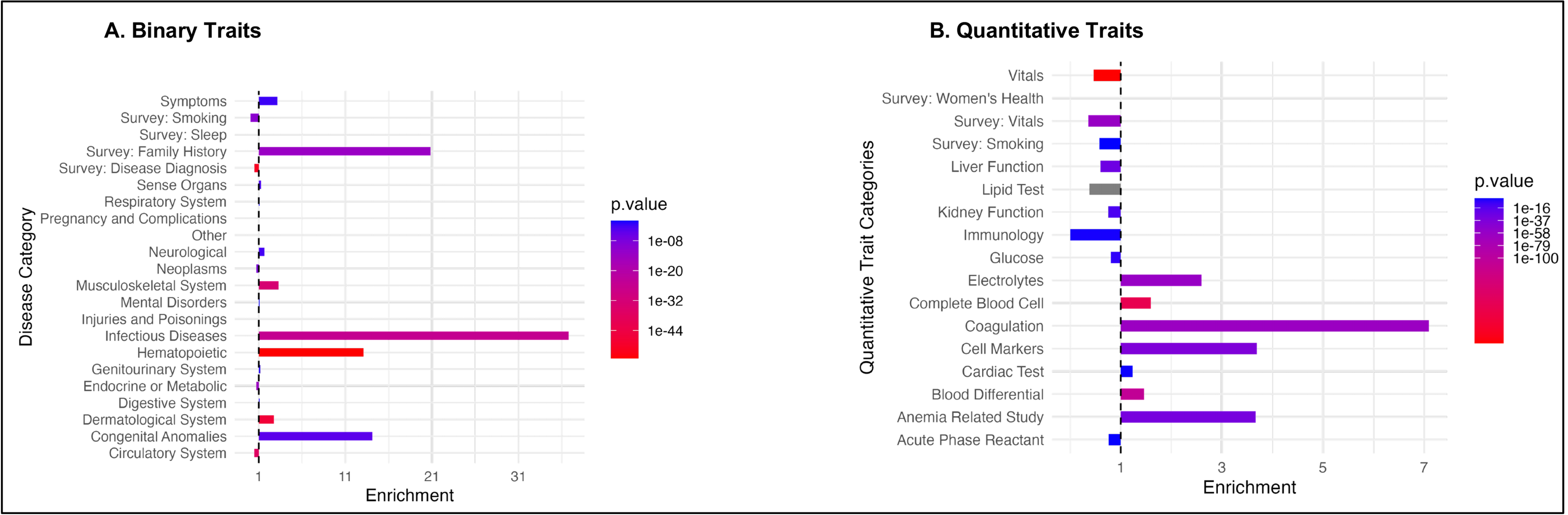

**Figure S3.** Enrichment analysis of novel associations across disease categories.

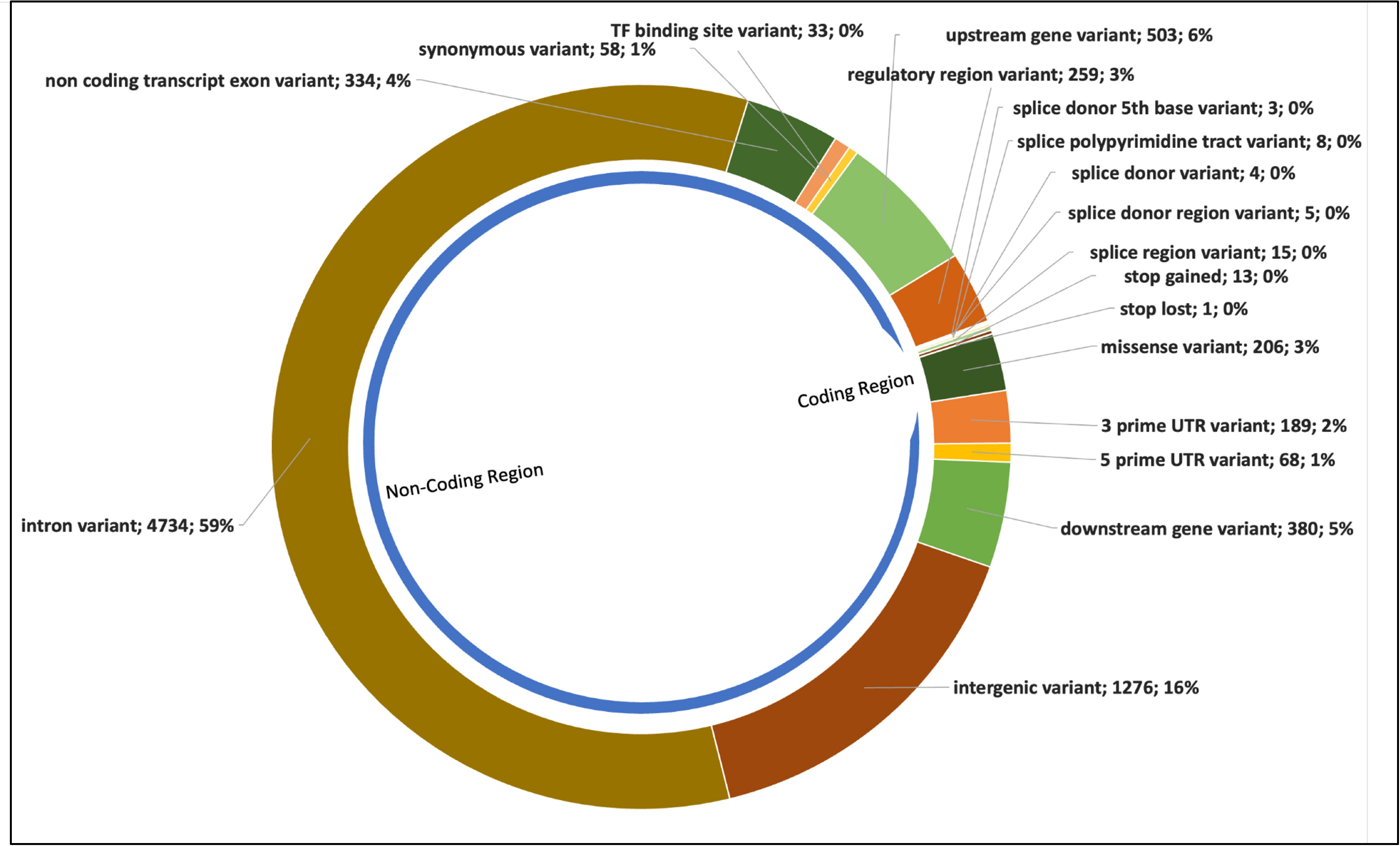

**Figure S4**. VEP annotations of lead SNPs identified from the meta-analysis.

**Figure S5.** Overview of Fine-Mapping Analysis and Results.

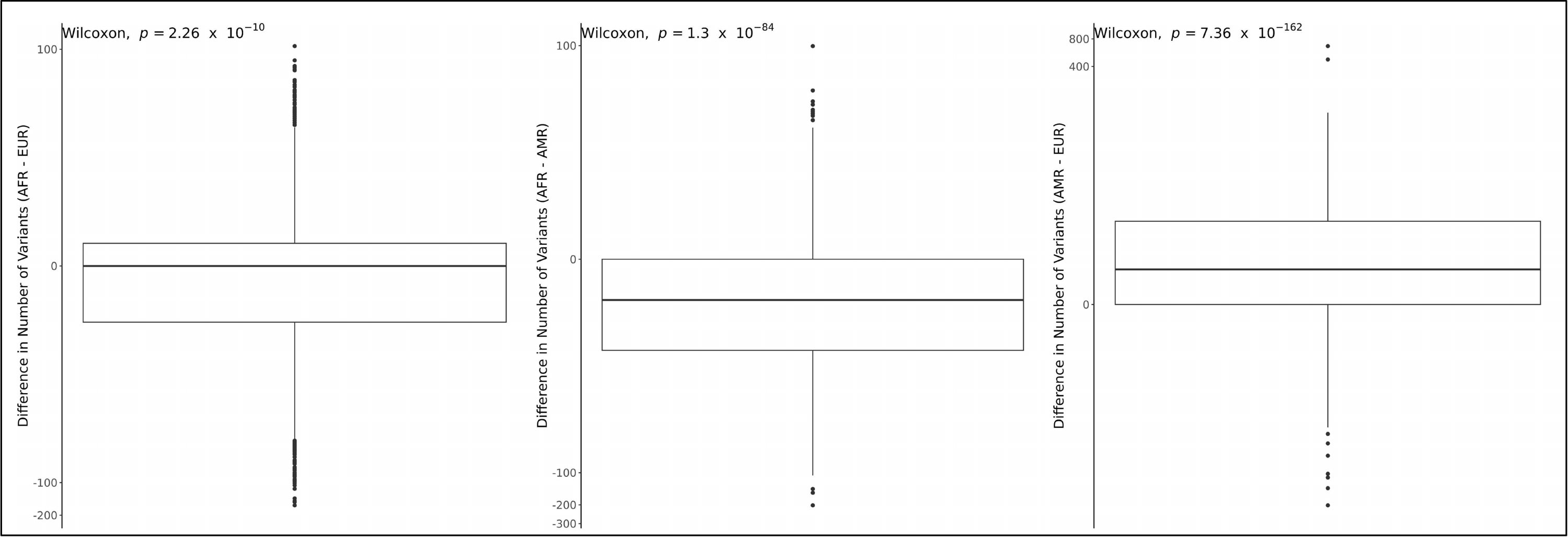
 **Figure S6**. Differences in credible set sizes for signals mapped in multiple populations.

**Figure S7.** Distributions of allele frequencies and RegulomeDB scores for fine-mapped variants.

**Figure S8.** Effect Sizes and allele frequencies for high-confidence and lead variants.

**Figure S9.** Cross-trait association of variants with two or more traits.

**Figure S10.** Heritability estimates of traits across four population groups.

**Figure S11A.** Locus Zoom plot of APOE signal. **Figure S11B**. Locus Zoom plot of APOE signal.

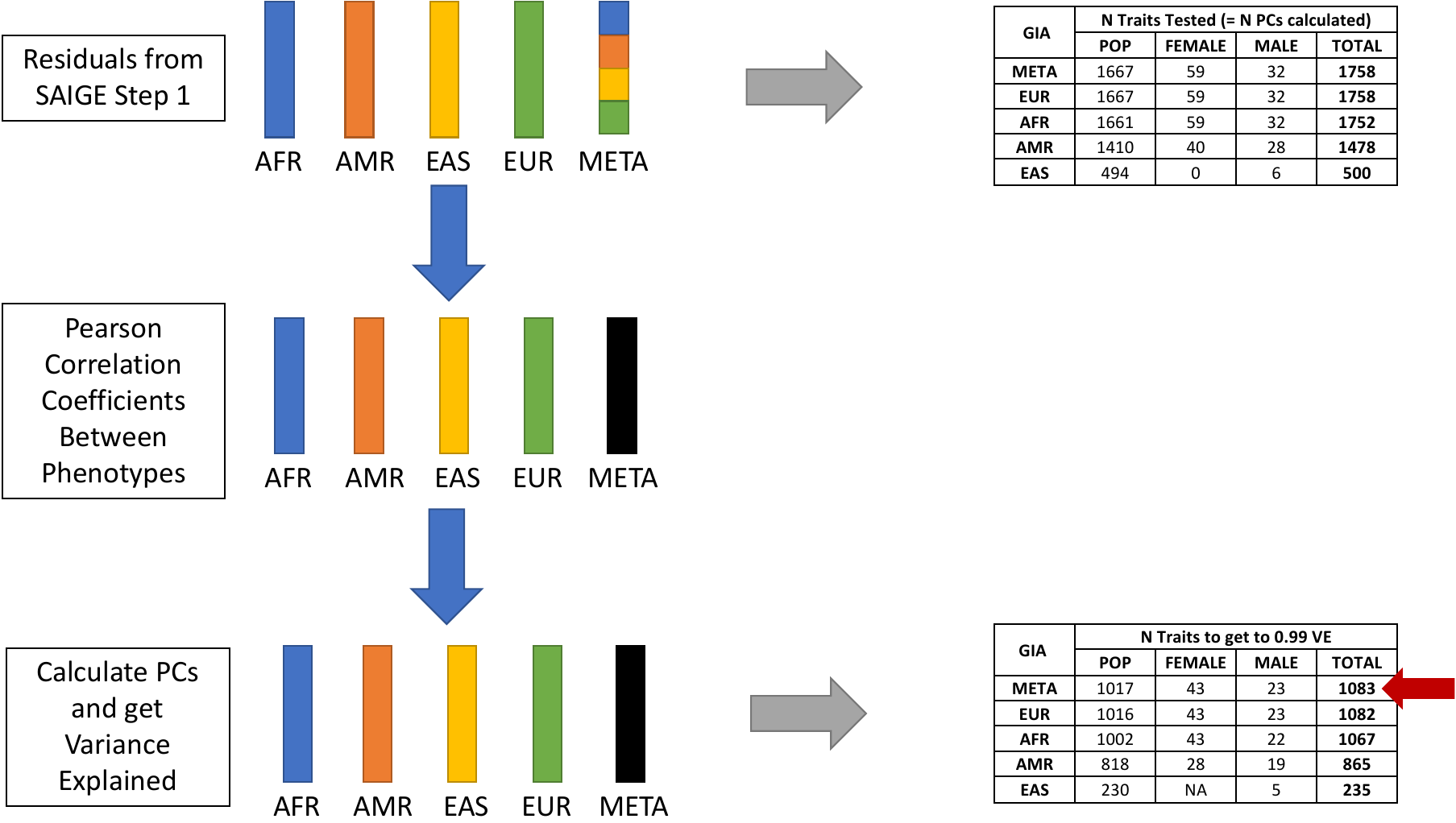

**Figure S12.** Flow diagram for determining the number of independent traits analyzed.

AFR = African ancestry, AMR = Admixed American ancestry, EAS = East Asian ancestry, EUR = European ancestry, META = all people included in the meta-analysis, GIA = genetically informed ancestry, POP = trait measured in everyone (ie lung cancer), FEMALE = trait only present in females (ie. ovarian cancer), MALE = trait present only in males (ie. prostate cancer), PC = principal component, VE = variance explained. Red arrow indicates the number used for Bonferroni correction.

**Figure S13. Principal component analysis.** The first ten principal components demonstrate how the MVP participants, represented in gray, are projected onto the 1000 genomes super populations. The final plot at the bottom shows a scree plot that indicates the variation of each principal component.

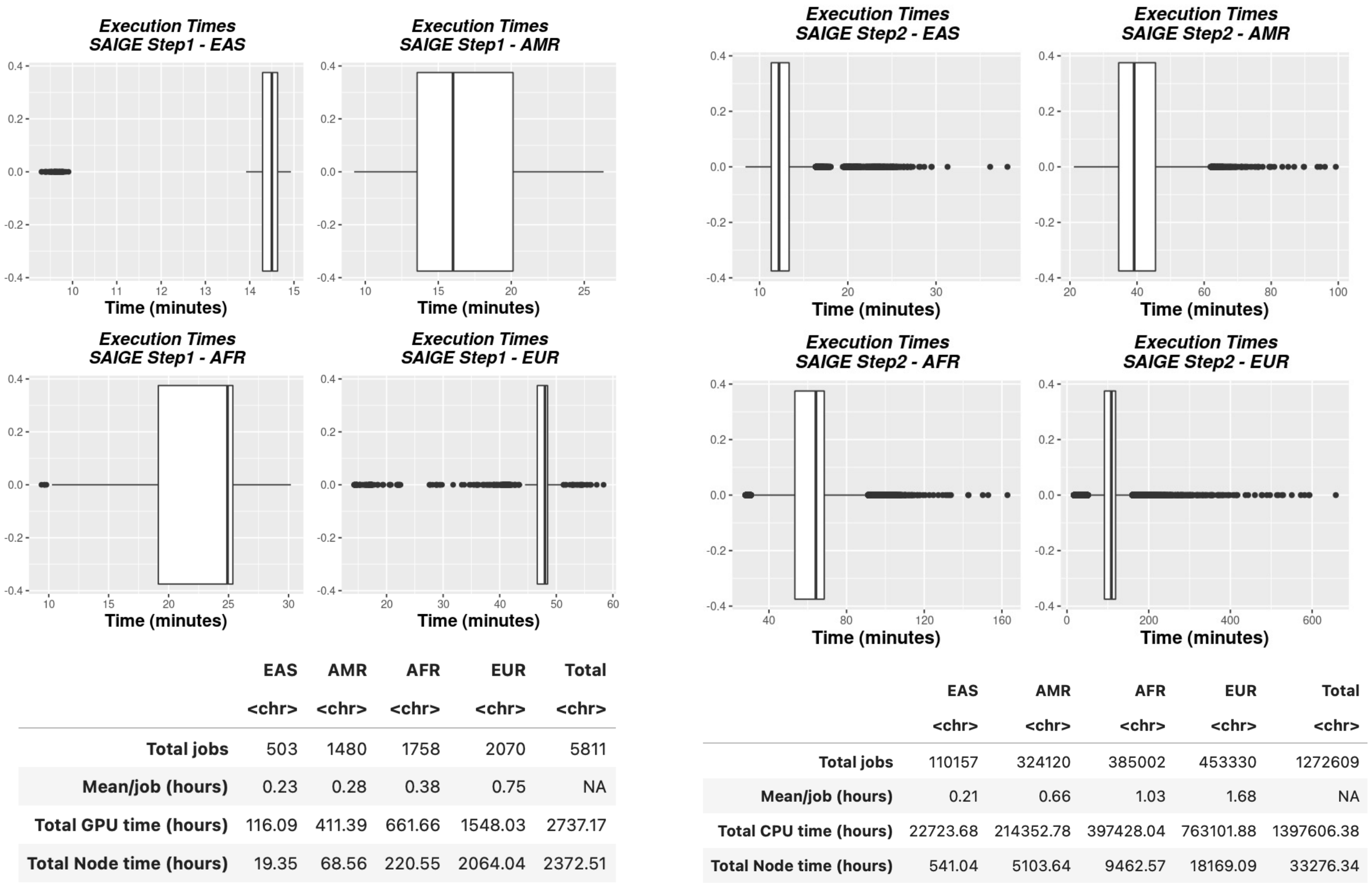

**Figure S14.** Execution times of optimized SAIGE on OLCF supercomputing infrastructure.

#### Contributions and Acknowledgments

**MVP Program Office**

- Sumitra Muralidhar, Ph.D., Program Director

US Department of Veterans Affairs, 810 Vermont Avenue NW, Washington, DC 20420

- Jennifer Moser, Ph.D., Associate Director, Scientific Programs

US Department of Veterans Affairs, 810 Vermont Avenue NW, Washington, DC 20420

- Jennifer E. Deen, B.S., Associate Director, Cohort & Public Relations

US Department of Veterans Affairs, 810 Vermont Avenue NW, Washington, DC 20420

**MVP Executive Committee**

- Co-Chair: Philip S. Tsao, Ph.D.

VA Palo Alto Health Care System, 3801 Miranda Avenue, Palo Alto, CA 94304

- Co-Chair: Sumitra Muralidhar, Ph.D.

US Department of Veterans Affairs, 810 Vermont Avenue NW, Washington, DC 20420

- J. Michael Gaziano, M.D., M.P.H.

VA Boston Healthcare System, 150 S. Huntington Avenue, Boston, MA 02130

- Elizabeth Hauser, Ph.D.

Durham VA Medical Center, 508 Fulton Street, Durham, NC 27705

- Amy Kilbourne, Ph.D., M.P.H.

VA HSR&D, 2215 Fuller Road, Ann Arbor, MI 48105

- Shiuh-Wen Luoh, M.D., Ph.D.

VA Portland Health Care System, 3710 SW US Veterans Hospital Rd, Portland, OR 97239

- Michael Matheny, M.D., M.S., M.P.H.

VA Tennessee Valley Healthcare System, 1310 24^th^ Ave. South, Nashville, TN 37212

- Dave Oslin, M.D.

Philadelphia VA Medical Center, 3900 Woodland Avenue, Philadelphia, PA 19104

**MVP Co-Principal Investigators**

- J. Michael Gaziano, M.D., M.P.H.

VA Boston Healthcare System, 150 S. Huntington Avenue, Boston, MA 02130

- Philip S. Tsao, Ph.D.

VA Palo Alto Health Care System, 3801 Miranda Avenue, Palo Alto, CA 94304

**MVP Core Operations**

- Lori Churby, B.S., Director, MVP Regulatory Affairs

VA Palo Alto Health Care System, 3801 Miranda Avenue, Palo Alto, CA 94304

- Stacey B. Whitbourne, Ph.D., Director, MVP Cohort Management

VA Boston Healthcare System, 150 S. Huntington Avenue, Boston, MA 02130

- Jessica V. Brewer, M.P.H., Director, MVP Recruitment & Enrollment

VA Boston Healthcare System, 150 S. Huntington Avenue, Boston, MA 02130

- Shahpoor (Alex) Shayan, M.S., Director, MVP Recruitment and Enrollment Informatics

VA Boston Healthcare System, 150 S. Huntington Avenue, Boston, MA 02130

- Luis E. Selva, Ph.D., Executive Director, MVP Biorepositories

VA Boston Healthcare System, 150 S. Huntington Avenue, Boston, MA 02130

- Saiju Pyarajan Ph.D., Director, Data and Computational Sciences

VA Boston Healthcare System, 150 S. Huntington Avenue, Boston, MA 02130

- Kelly Cho, M.P.H, Ph.D., Director, MVP Phenomics Data Core

VA Boston Healthcare System, 150 S. Huntington Avenue, Boston, MA 02130

- Scott L. DuVall, Ph.D., Director, VA Informatics and Computing Infrastructure (VINCI)

VA Salt Lake City Health Care System, 500 Foothill Drive, Salt Lake City, UT 84148

- Mary T. Brophy M.D., M.P.H., Director, VA Central Biorepository

VA Boston Healthcare System, 150 S. Huntington Avenue, Boston, MA 02130

- MVP Coordinating Centers
  - MVP Coordinating Center, Boston - J. Michael Gaziano, M.D., M.P.H.

VA Boston Healthcare System, 150 S. Huntington Avenue, Boston, MA 02130

- MVP Coordinating Center, Palo Alto – Philip S. Tsao, Ph.D.

VA Palo Alto Health Care System, 3801 Miranda Avenue, Palo Alto, CA 94304

- MVP Information Center, Canandaigua – Brady Stephens, M.S.

Canandaigua VA Medical Center, 400 Fort Hill Avenue, Canandaigua, NY 14424

- Cooperative Studies Program Clinical Research Pharmacy Coordinating Center, Albuquerque – Todd Connor, Pharm.D.; Dean P. Argyres, B.S., M.S.

New Mexico VA Health Care System, 1501 San Pedro Drive SE, Albuquerque, NM 87108

**MVP Publications and Presentations Committee**

- Co-Chair: Themistocles L. Assimes, M.D., Ph. D

VA Palo Alto Health Care System, 3801 Miranda Avenue, Palo Alto, CA 94304

- Co-Chair: Adriana Hung, M.D.; M.P.H

VA Tennessee Valley Healthcare System, 1310 24^th^ Ave. South, Nashville, TN 37212

- Co-Chair: Henry Kranzler, M.D.

Philadelphia VA Medical Center, 3900 Woodland Avenue, Philadelphia, PA 19104

**MVP Local Site Investigators**

- Samuel Aguayo, M.D., Phoenix VA Health Care System

650 E. Indian School Road, Phoenix, AZ 85012

- Sunil Ahuja, M.D., South Texas Veterans Health Care System

7400 Merton Minter Boulevard, San Antonio, TX 78229

- Kathrina Alexander, M.D., Veterans Health Care System of the Ozarks

1100 North College Avenue, Fayetteville, AR 72703

- Xiao M. Androulakis, M.D., Columbia VA Health Care System

6439 Garners Ferry Road, Columbia, SC 29209

- Prakash Balasubramanian, M.D., William S. Middleton Memorial Veterans Hospital

2500 Overlook Terrace, Madison, WI 53705

- Zuhair Ballas, M.D., Iowa City VA Health Care System

601 Highway 6 West, Iowa City, IA 52246-2208

- Jean Beckham, Ph.D., Durham VA Medical Center

508 Fulton Street, Durham, NC 27705

- Sujata Bhushan, M.D., VA North Texas Health Care System

4500 S. Lancaster Road, Dallas, TX 75216

- Edward Boyko, M.D., VA Puget Sound Health Care System

1660 S. Columbian Way, Seattle, WA 98108-1597

- David Cohen, M.D., Portland VA Medical Center

3710 SW U.S. Veterans Hospital Road, Portland, OR 97239

- Louis Dellitalia, M.D., Birmingham VA Medical Center

700 S. 19th Street, Birmingham AL 35233

- L. Christine Faulk, M.D., Robert J. Dole VA Medical Center

5500 East Kellogg Drive, Wichita, KS 67218-1607

- Joseph Fayad, M.D., VA Southern Nevada Healthcare System

6900 North Pecos Road, North Las Vegas, NV 89086

- Daryl Fujii, Ph.D., VA Pacific Islands Health Care System

459 Patterson Rd, Honolulu, HI 96819

- Saib Gappy, M.D., John D. Dingell VA Medical Center

4646 John R Street, Detroit, MI 48201

- Frank Gesek, Ph.D., White River Junction VA Medical Center

163 Veterans Drive, White River Junction, VT 05009

- Jennifer Greco, M.D., Sioux Falls VA Health Care System

2501 W 22nd Street, Sioux Falls, SD 57105

- Michael Godschalk, M.D., Richmond VA Medical Center

1201 Broad Rock Blvd., Richmond, VA 23249

- Todd W. Gress, M.D., Ph.D., Hershel “Woody” Williams VA Medical Center

1540 Spring Valley Drive, Huntington, WV 25704

- Samir Gupta, M.D., M.S.C.S., VA San Diego Healthcare System

3350 La Jolla Village Drive, San Diego, CA 92161

- Salvador Gutierrez, M.D., Edward Hines, Jr. VA Medical Center

5000 South 5th Avenue, Hines, IL 60141

- John Harley, M.D., Ph.D., Cincinnati VA Medical Center

3200 Vine Street, Cincinnati, OH 45220

- Kimberly Hammer, Ph.D., Fargo VA Health Care System

2101 N. Elm, Fargo, ND 58102

- Mark Hamner, M.D., Ralph H. Johnson VA Medical Center

109 Bee Street, Mental Health Research, Charleston, SC 29401

- Adriana Hung, M.D., M.P.H., VA Tennessee Valley Healthcare System

1310 24th Avenue, South Nashville, TN 37212

- Robin Hurley, M.D., W.G. (Bill) Hefner VA Medical Center

1601 Brenner Ave, Salisbury, NC 28144

- Pran Iruvanti, D.O., Ph.D., Hampton VA Medical Center

100 Emancipation Drive, Hampton, VA 23667

- Frank Jacono, M.D., VA Northeast Ohio Healthcare System

10701 East Boulevard, Cleveland, OH 44106

- Darshana Jhala, M.D., Philadelphia VA Medical Center

3900 Woodland Avenue, Philadelphia, PA 19104

- Scott Kinlay, M.B.B.S., Ph.D., VA Boston Healthcare System

150 S. Huntington Avenue, Boston, MA 02130

- Jon Klein, M.D., Ph.D., Louisville VA Medical Center

800 Zorn Avenue, Louisville, KY 40206

- Michael Landry, Ph.D., Southeast Louisiana Veterans Health Care System

2400 Canal Street, New Orleans, LA 70119

- Peter Liang, M.D., M.P.H., VA New York Harbor Healthcare System

423 East 23rd Street, New York, NY 10010

- Suthat Liangpunsakul, M.D., M.P.H., Richard Roudebush VA Medical Center

1481 West 10th Street, Indianapolis, IN 46202

- Jack Lichy, M.D., Ph.D., Washington DC VA Medical Center

50 Irving St, Washington, D. C. 20422

- C. Scott Mahan, M.D., Charles George VA Medical Center

1100 Tunnel Road, Asheville, NC 28805

- Ronnie Marrache, M.D., VA Maine Healthcare System

1 VA Center, Augusta, ME 04330

- Stephen Mastorides, M.D., James A. Haley Veterans’ Hospital

13000 Bruce B. Downs Blvd, Tampa, FL 33612

- Elisabeth Mates M.D., Ph.D., VA Sierra Nevada Health Care System

975 Kirman Avenue, Reno, NV 89502

- Kristin Mattocks, Ph.D., M.P.H., Central Western Massachusetts Healthcare System

421 North Main Street, Leeds, MA 01053

- Paul Meyer, M.D., Ph.D., Southern Arizona VA Health Care System

3601 S 6th Avenue, Tucson, AZ 85723

- Jonathan Moorman, M.D., Ph.D., James H. Quillen VA Medical Center

Corner of Lamont & Veterans Way, Mountain Home, TN 37684

- Timothy Morgan, M.D., VA Long Beach Healthcare System

5901 East 7th Street Long Beach, CA 90822

- Maureen Murdoch, M.D., M.P.H., Minneapolis VA Health Care System

One Veterans Drive, Minneapolis, MN 55417

- James Norton, Ph.D., VA Health Care Upstate New York

113 Holland Avenue, Albany, NY 12208

- Olaoluwa Okusaga, M.D., Michael E. DeBakey VA Medical Center

2002 Holcombe Blvd, Houston, TX 77030

- Kris Ann Oursler, M.D., Salem VA Medical Center

1970 Roanoke Blvd, Salem, VA 24153

- Ana Palacio, M.D., M.P.H., Miami VA Health Care System

1201 NW 16th Street, 11 GRC, Miami FL 33125

- Samuel Poon, M.D., Manchester VA Medical Center

718 Smyth Road, Manchester, NH 03104

- Emily Potter, Pharm.D., VA Eastern Kansas Health Care System

4101 S 4th Street Trafficway, Leavenworth, KS 66048

- Michael Rauchman, M.D., St. Louis VA Health Care System

915 North Grand Blvd, St. Louis, MO 63106

- Richard Servatius, Ph.D., Syracuse VA Medical Center

800 Irving Avenue, Syracuse, NY 13210

- Satish Sharma, M.D., Providence VA Medical Center

830 Chalkstone Avenue, Providence, RI 02908

- River Smith, Ph.D., Eastern Oklahoma VA Health Care System

1011 Honor Heights Drive, Muskogee, OK 74401

- Peruvemba Sriram, M.D., N. FL/S. GA Veterans Health System

1601 SW Archer Road, Gainesville, FL 32608

- Patrick Strollo, Jr., M.D., VA Pittsburgh Health Care System

University Drive, Pittsburgh, PA 15240

- Neeraj Tandon, M.D., Overton Brooks VA Medical Center

510 East Stoner Ave, Shreveport, LA 71101

- Philip Tsao, Ph.D., VA Palo Alto Health Care System

3801 Miranda Avenue, Palo Alto, CA 94304-1290

- Gerardo Villareal, M.D., New Mexico VA Health Care System

1501 San Pedro Drive, S.E. Albuquerque, NM 87108

- Agnes Wallbom, M.D., M.S., VA Greater Los Angeles Health Care System

11301 Wilshire Blvd, Los Angeles, CA 90073

- Jessica Walsh, M.D., VA Salt Lake City Health Care System

500 Foothill Drive, Salt Lake City, UT 84148

- John Wells, Ph.D., Edith Nourse Rogers Memorial Veterans Hospital

200 Springs Road, Bedford, MA 01730

- Jeffrey Whittle, M.D., M.P.H., Clement J. Zablocki VA Medical Center

5000 West National Avenue, Milwaukee, WI 53295

- Mary Whooley, M.D., San Francisco VA Health Care System

4150 Clement Street, San Francisco, CA 94121

- Allison E. Williams, N.D., Ph.D., R.N, Bay Pines VA Healthcare System

10,000 Bay Pines Blvd Bay Pines, FL 33744

- Peter Wilson, M.D., Atlanta VA Medical Center

1670 Clairmont Road, Decatur, GA 30033

- Junzhe Xu, M.D., VA Western New York Healthcare System

3495 Bailey Avenue, Buffalo, NY 14215-1199

- Shing Shing Yeh, Ph.D., M.D., Northport VA Medical Center

79 Middleville Road, Northport, NY 11768

#### References

1.	A. Price, *EIGENSOFT [Internet]. Available from* (http://www.hsph.harvard.edu/alkes-price/software/.).

2.	Y. Deng, W. Pan, Improved Use of Small Reference Panels for Conditional and Joint Analysis with GWAS Summary Statistics. *Genetics*. **209**, 401–408 (2018).

3.	R. E. Peterson, K. Kuchenbaecker, R. K. Walters, C.-Y. Chen, A. B. Popejoy, S. Periyasamy, M. Lam, C. Iyegbe, R. J. Strawbridge, L. Brick, C. E. Carey, A. R. Martin, J. L. Meyers, J. Su, J. Chen, A. C. Edwards, A. Kalungi, N. Koen, L. Majara, E. Schwarz, J. W. Smoller, E. A. Stahl, P. F. Sullivan, E. Vassos, B. Mowry, M. L. Prieto, A. Cuellar-Barboza, T. B. Bigdeli, H. J. Edenberg, H. Huang, L. E. Duncan, Genome-wide Association Studies in Ancestrally Diverse Populations: Opportunities, Methods, Pitfalls, and Recommendations. *Cell*. **179**, 589–603 (2019).

4.	C. C. Chang, C. C. Chow, L. C. Tellier, S. Vattikuti, S. M. Purcell, J. J. Lee, Second-generation PLINK: rising to the challenge of larger and richer datasets. *GigaScience*. **4** (2015), doi:10.1186/s13742-015-0047-8.

5.	M. Kanai, J. C. Ulirsch, J. Karjalainen, M. Kurki, K. J. Karczewski, E. Fauman, Q. S. Wang, H. Jacobs, F. Aguet, K. G. Ardlie, N. Kerimov, K. Alasoo, C. Benner, K. Ishigaki, S. Sakaue, S. Reilly, The BioBank Japan Project, FinnGen, Y. Kamatani, K. Matsuda, A. Palotie, B. M. Neale, R. Tewhey, P. C. Sabeti, Y. Okada, M. J. Daly, H. K. Finucane, “Insights from complex trait fine-mapping across diverse populations” (preprint, Genetic and Genomic Medicine, 2021), , doi:10.1101/2021.09.03.21262975.

6.	O. Weissbrod, F. Hormozdiari, C. Benner, R. Cui, J. Ulirsch, S. Gazal, A. P. Schoech, B. van de Geijn, Y. Reshef, C. Márquez-Luna, L. O’Connor, M. Pirinen, H. K. Finucane, A. L. Price, Functionally informed fine-mapping and polygenic localization of complex trait heritability. *Nat. Genet.* **52**, 1355–1363 (2020).

7.	K. Yuan, R. J. Longchamps, A. F. Pardiñas, M. Yu, T.-T. Chen, S.-C. Lin, Y. Chen, M. Lam, R. Liu, Y. Xia, Z. Guo, W. Shi, C. Shen, The Schizophrenia Workgroup of Psychiatric Genomics Consortium, M. J. Daly, B. Neale, Y.-C. A. Feng, Y.-F. Lin, C.-Y. Chen, M. O’Donovan, T. Ge, H. Huang, “Fine-mapping across diverse ancestries drives the discovery of putative causal variants underlying human complex traits and diseases” (preprint, Genetic and Genomic Medicine, 2023), , doi:10.1101/2023.01.07.23284293.

8.	M.-H. Chen, L. M. Raffield, A. Mousas, S. Sakaue, J. E. Huffman, A. Moscati, B. Trivedi, T. Jiang, P. Akbari, D. Vuckovic, E. L. Bao, X. Zhong, R. Manansala, V. Laplante, M. Chen, K. S. Lo, H. Qian, C. A. Lareau, M. Beaudoin, K. A. Hunt, M. Akiyama, T. M. Bartz, Y. Ben-Shlomo, A. Beswick, J. Bork-Jensen, E. P. Bottinger, J. A. Brody, F. J. A. van Rooij, K. Chitrala, K. Cho, H. Choquet, A. Correa, J. Danesh, E. Di Angelantonio, N. Dimou, J. Ding, P. Elliott, T. Esko, M. K. Evans, J. S. Floyd, L. Broer, N. Grarup, M. H. Guo, A. Greinacher, J. Haessler, T. Hansen, J. M. M. Howson, Q. Q. Huang, W. Huang, E. Jorgenson, T. Kacprowski, M. Kähönen, Y. Kamatani, M. Kanai, S. Karthikeyan, F. Koskeridis, L. A. Lange, T. Lehtimäki, M. M. Lerch, A. Linneberg, Y. Liu, L.-P. Lyytikäinen, A. Manichaikul, H. C. Martin, K. Matsuda, K. L. Mohlke, N. Mononen, Y. Murakami, G. N. Nadkarni, M. Nauck, K. Nikus, W. H. Ouwehand, N. Pankratz, O. Pedersen, M. Preuss, B. M. Psaty, O. T. Raitakari, D. J. Roberts, S. S. Rich, B. A. T. Rodriguez, J. D. Rosen, J. I. Rotter, P. Schubert, C. N. Spracklen, P. Surendran, H. Tang, J.-C. Tardif, R. C. Trembath, M. Ghanbari, U. Völker, H. Völzke, N. A. Watkins, A. B. Zonderman, VA Million Veteran Program, P. W. F. Wilson, Y. Li, A. S. Butterworth, J.-F. Gauchat, C. W. K. Chiang, B. Li, R. J. F. Loos, W. J. Astle, E. Evangelou, D. A. van Heel, V. G. Sankaran, Y. Okada, N. Soranzo, A. D. Johnson, A. P. Reiner, P. L. Auer, G. Lettre, Trans-ethnic and Ancestry-Specific Blood-Cell Genetics in 746,667 Individuals from 5 Global Populations. *Cell*. **182**, 1198-1213.e14 (2020).

9.	A. Li, R. F. Barber, Multiple Testing with the Structure-Adaptive Benjamini–Hochberg Algorithm. *J. R. Stat. Soc. Ser. B Stat. Methodol.* **81**, 45–74 (2019).

10.	T. T. Cai, W. Sun, Y. Xia, LAWS: A Locally Adaptive Weighting and Screening Approach to Spatial Multiple Testing. *J. Am. Stat. Assoc.* **117**, 1370–1383 (2022).

11.	Y. Benjamini, Y. Hochberg, Controlling the False Discovery Rate: A Practical and Powerful Approach to Multiple Testing. *J. R. Stat. Soc. Ser. B Methodol.* **57**, 289–300 (1995).

12.	M. Liu, E. Katsevich, L. Janson, A. Ramdas, Fast and Powerful Conditional Randomization Testing via Distillation (2021), , doi:10.48550/arXiv.2006.03980.
